# Locally adaptive conformal prediction intervals for polygenic score-based phenotype prediction via residual normalization and data-driven stratification

**DOI:** 10.64898/2026.05.28.26354326

**Authors:** Yu Yun, Xingjie Hao, Yan Dora Zhang

## Abstract

Quantifying uncertainty in polygenic score (PGS)-based phenotype prediction is crucial for the integration of genomic data into precision medicine. While the PGS provides a fundamental pivot for point estimation, clinical decision-making necessitates the construction of well-calibrated prediction intervals that reliably encompass the true phenotypic values. However, phenotypic residuals are frequently characterized by complex heteroscedasticity and stratified variance structures across diverse demographic contexts. Existing approaches often rely on global calibration mechanisms, which fail to account for such localized variance structures and lead to systematic miscalibration within specific subpopulations. To bridge this gap, we propose Clustering-based Split Conformal Prediction with Normalized Residuals (C-SCNR), a versatile framework based on Split Conformal Prediction. By adopting residual normalization and incorporating a repetitive ‘split-and-cluster’ mechanism, C-SCNR dynamically identifies latent error strata and applies fine-grained adjustments to the resulting intervals.

Our framework requires no distributional assumptions regarding the phenotype, is compatible with any PGS method, and flexibly accommodates biologically-informed grouping. Simulation studies demonstrate that our framework consistently outperforms existing methods across diverse error distributions. In real-data applications analyzing Body mass index (BMI), Low-density lipoprotein (LDL) cholesterol, and High-density lipoprotein (HDL) cholesterol in the UK Biobank, C-SCNR effectively resolves the coverage deficiencies of existing methods in specific subgroups and consistently yields superior localized calibration. Overall, C-SCNR represents a flexible and powerful framework for constructing high-resolution context-specific prediction intervals, thereby facilitating more reliable clinical interpretations of polygenic risk.

## 1 Introduction

The advancement of precision medicine relies on the ability to accurately assess an individual’s risk of developing complex diseases and predict phenotypic outcomes^1–3^. Over the past two decades, genome-wide association studies (GWAS) have revolutionized our understanding of the human genetic architecture by identifying millions of single-nucleotide polymorphisms (SNPs) associated with complex disorders^4–14^. However, since the majority of these variants exert only marginal effects, they often fail to provide clinical insights when considered in isolation. This limitation has led to the rise of the polygenic score (PGS), or polygenic risk score (PRS), which aggregates the cumulative effects of thousands of genetic variants into a weighted measure of genetic susceptibility. The PGS provides a quantitative summary of an individual’s genetic liability with respect to a phenotype, and its predictive value has been extensively validated across a broad spectrum of complex diseases and traits^6–8,10–15^. Consequently, the PGS has transitioned from a research tool to a potential clinical instrument, supporting the shift towards more proactive disease detection, prevention, and therapeutic intervention^7,8,10–12,14,15^.

Despite the predictive power of the PGS, the transition from genetic risk estimation to accurate phenotypic prediction remains a formidable statistical challenge. Various statistical methods have been developed to construct PGSs, primarily focusing on optimizing the estimation of SNP effect sizes^24–29^. However, the resulting scores are inevitably subject to estimation errors stemming from limited sample sizes, linkage disequilibrium (LD), and misspecified models. Moreover, in clinical practice, the PGS is typically utilized as a predictor of the target trait within a regression framework, integrated with additional demographic covariates^30^. Such regression models are inherently imperfect, failing to capture the full range of uncertainty underlying complex traits. Therefore, reliance on point estimates alone is insufficient for informed clinical decision-making. To properly characterize the true phenotypic values of target individuals, it is imperative to move beyond point estimates and rigorously quantify the prediction uncertainty by constructing well-calibrated prediction intervals. To address the need for uncertainty quantification in phenotype prediction, several statistical frameworks have recently emerged. One such approach, PredInterval ^33^, adopts a non-parametric framework that leverages the quantile information of phenotypic residuals through cross-validation to construct intervals. On the other hand, CalPred^34^ has been developed to address the challenge of context-specific prediction by incorporating diverse contextual information, such as age, sex, and socioeconomic status, into the calibration model, which allows for the construction of prediction intervals that adapt to the specific background of each individual.

The clinical utility of a risk model is fundamentally tied to its ability to perform risk stratification, categorizing individuals into meaningful subgroups based on distinct risk profiles^12,13,15,16,22^. However, current state-of-the-art frameworks fall short in maintaining uniform calibration across these diverse strata. PredInterval, for instance, operates under a global quantile calibration, which may fail to capture the localized shifts in error scales across different demographic subgroups. This reliance on a global quantile of absolute residuals likely yields constant-width intervals and therefore fails to adapt to heteroscedastic error structures. Similarly, CalPred relies on specific distributional and contextual assumptions. Such assumptions may not hold when phenotypic error distributions exhibit underlying discontinuities driven by latent demographic and environmental factors. Meanwhile, this challenge is exacerbated by the fact that the genetic architecture of complex traits is rarely static. Extensive evidence suggests that for many complex traits, such as Body mass index (BMI), lipid levels, and blood pressure, the underlying genetic and environmental effects are strongly modulated by demographic factors including age and sex^17–23^. The predictive power of the PGS often fluctuates across age intervals and common disorders frequently exhibit sex-dimorphism in prevalence and etiology. Consequently, phenotypic error distributions are not uniform but are instead subjected to stratified variance, as a result of these complex interactions. Under stratified structures, the existing models are prone to localized miscalibration, where prediction intervals become over-conservative for certain subgroups while providing insufficient coverage for others. Such disparities in calibration undermine the integrity of risk stratification, leading to inappropriate clinical decisions for individuals residing in the under-covered groups.

To address these challenges, we introduce Clustering-based Split Conformal Prediction with Normalized Residuals (C-SCNR), a robust and granular framework designed for the heterogeneous error landscapes of complex phenotypes. Motivated by the Conformal Prediction framework^31,32^, our approach is based on a two-stage methodological progression, extending from a generalized residual normalization strategy to a dynamic, clustering-based aggregation framework. First, we propose Split Conformal Prediction with Normalized Residuals (SCNR), which utilizes an estimated variance function to normalize the phenotypic residuals. By modeling the conditional variance as a function of relevant covariates, such as sex, age, and genetic ancestry, SCNR accounts for the broad heteroscedasticity observed in complex traits. This residual normalization ensures that the underlying error distribution is more homogenized, providing a foundation for the subsequent conformal prediction. However, recognizing that complex human traits often exhibit stratified variance structures that cannot be fully captured by continuous variance models, we extend this framework to Clustering-based SCNR (C-SCNR). C-SCNR incorporates a ‘split-and-cluster’ mechanism, where the calibration data is iteratively partitioned and clustered to identify latent, homogeneous error strata. By aggregating the calibration results across repetitions, C-SCNR achieves more refined localized validity. C-SCNR is model-agnostic, requiring no specific distributional assumptions regarding the phenotype. Moreover, it accommodates flexible user-specified biologically motivated groupings and allows for data-driven stratification.

To evaluate the performance of our proposed framework, we conducted comprehensive simulation studies. These simulations demonstrate that while the global normalization strategy SCNR is designed to account for broad continuous heteroscedasticity through a fitted variance function, it encounters limitations in the presence of stratified error structures. In contrast, C-SCNR consistently resolves the localized calibration gaps while retaining the advantages of SCNR, thereby maintaining the target coverage across diverse phenotypic distributions. We also applied our method and the existing approaches to analyze several quantitative traits in the UK Biobank, including Body mass index (BMI), Low-density lipoprotein (LDL) cholesterol, and High-density lipoprotein (HDL) cholesterol. For our primary configuration, we selected age and sex as the base grouping variables. This specific configuration is motivated by their well-documented age-dependent and sex-dimorphic genetic architectures^17,21,22^. However, we emphasize that this manual grouping serves as a foundational heuristic rather than a rigid optimum. The most effective grouping is specific to the phenotype under study and should remain flexible to its particular architecture. The results illustrate that C-SCNR consistently outperforms other frameworks by providing high-resolution, context-specific prediction intervals that remain valid across detailed demographic backgrounds. Overall, this study provides a versatile framework for locally-calibrated uncertainty quantification in PGS-based phenotype prediction, bridging the gap between theoretical conformal prediction and practical needs of clinical practice.

## 2 Methods and Materials

### Prediction Interval Construction with Conformal Prediction

We first frame the construction of PGS-based prediction intervals for quantitative phenotypes within the Conformal Prediction framework ^32^. Consider a dataset containing *N* individuals, where *Y*_*i*_ denotes the phenotypic value and ***G***_*i*_ ∈ {0, 1, 2}^*M*^ represents the genotype vector across *M* SNPs for the *i*-th individual, *i* ∈ {1,…, *N*}. For any given PGS method, let 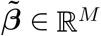 denote the vector of trained PGS weights. The corresponding PGS is calculated as 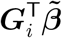 for each individual. To account for population structure and demographic factors, the PGS is treated as a predictor of the target trait and integrated with other appropriate covariates to obtain the predicted phenotypic value *Ŷ*_*i*_, typically through a regression model^30^. Let 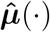 denote an estimator of the underlying regression function such that 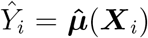, where ***X***_*i*_ is the vector of predictors for individual *i*, including the PGS and other covariates. For a new target individual *N* + 1 with predictors ***X***_*N*+1_, the Conformal Prediction addresses the prediction interval construction problem non-parametrically. A naive prediction interval for the phenotypic value *Y*_*N*+1_ at a target coverage level of 1 − *α* can be constructed as:

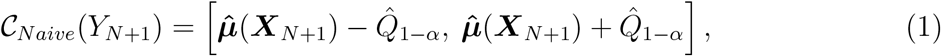

where 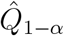 represents the (1 − *α*)-quantile of the empirical distribution of absolute residuals *R*_*i*_ = |*Y*_*i*_ − *Ŷ*_*i*_|, *i* ∈ {1,…, *N*}. Here, the absolute residual serves as a form of the conformity score function in the context of conformal inference. In fact, it can be replaced by any function that is symmetric with a decreasing density on [0, ∞). Since the residuals are evaluated on the same data used to fit 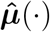, the empirical distribution of these in-sample residuals is systematically biased downward. Consequently, such naive prediction intervals typically suffer from under-coverage.

To address the downward bias in the coverage of naive intervals, the Full Conformal Prediction adopts a strategy based on augmented regression. Specifically, it fits the model using the original training data combined with each target sample ***X***_*N*+1_ paired with a predefined set of trial values 𝒴_*trial*_, one at a time. In other words, for a target dataset of *N*_*test*_ individuals, the number of augmented regressions to be fitted is *O*(*N*_*test*_ · |𝒴_*trial*_|). Although theoretically guaranteed to provide valid coverage, such a strategy presents limitations in terms of computational complexity and practicality.

To strike a balance between statistical efficiency and computational efficiency, popular alternative approaches include Jackknife+, CV+, and Split Conformal Prediction^31^. For instance, PredInterval adopted the CV+ framework to construct prediction intervals through *K*-fold cross-validation^33^. Under this framework, the training dataset is partitioned into *K* disjoint subsets *S*_1_,…, *S*_*K*_ of equal size. For individuals in each subset, their PGSs are calculated using the PGS weights obtained by treating the remaining *K* − 1 subsets as training data. Let *S*_*k*(*i*)_ denote the subset containing individual *i*, where *k*(*i*) ∈ {1,…, *K*}. The predicted phenotypic value is given by 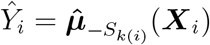, where 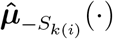 is an estimator of the regression function trained using the training data with the subset *S*_*k*(*i*)_ removed. The resulting absolute residuals are 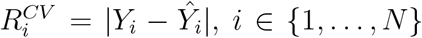. For a target individual, *K* distinct predicted values are calculated, and the absolute residuals corresponding to each subset *S*_*k*_, *k* ∈ {1,…, *K*} are aggregated to form the sets of possible lower and upper phenotypic bounds. The resulting 1 − *α* prediction interval is defined as:

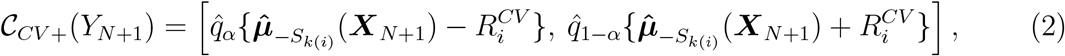

where 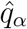denotes the *α*-th quantile, and *i* ∈ {1,…, *N*}. Notably, the CV+ framework is an extension of Jackknife+, where the number of folds *K* = *N* for Jackknife+.

In contrast, Split Conformal Prediction randomly splits the training dataset into two non-overlapping parts: the proper training set 𝒟_1_ and the calibration set 𝒟_2_, with sample sizes *N*_*train*_ and *N*_*cal*_, respectively. The regression function is fitted exclusively on the proper training set 𝒟_1_, yielding the regression function estimator 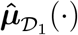. The absolute residuals are then evaluated on the calibration set as 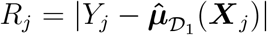, *j* ∈ 𝒟_2_. Let 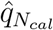 denote the ⌈(1 − *α*)(*N*_*cal*_ + 1)⌉-th smallest value in {*R*_*j*_: *j* ∈ 𝒟_2_}. The prediction interval via Split Conformal Prediction is built as:

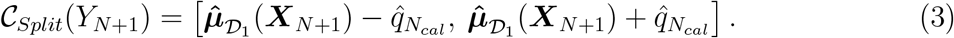

In terms of computational complexity, CV+ requires multiple model fits, while Split Conformal Prediction requires one model fitting only. Moreover, the fundamental difference between these two methods is that CV+ leverages all available data for both model fitting and calibration, whereas Split Conformal Prediction strictly segregates these tasks into two disjoint subsets. However, their shared reliance on a global quantile of absolute residuals implicitly assumes a homoscedastic error structure, resulting in prediction intervals with roughly constant widths over the target data. Hence, a more flexible calibration strategy capable of capturing the underlying heteroscedastic structure is required, as discussed in the following section.

### Overview of SCNR and C-SCNR

To address the aforementioned limitations and enhance the local adaptivity of the resulting prediction intervals, we propose adopting the locally-weighted Split Conformal Prediction framework, which accounts for individual-level heteroscedasticity.

In accordance with the principles of conformal inference, the conformity score function can be generalized to a normalized version, where the original residuals are scaled inversely by an estimated local spread of the errors. Based on the Split Conformal Prediction framework, the normalized absolute residuals are defined as:

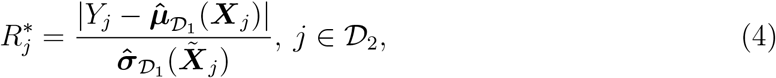

where 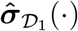 denotes an estimator of the conditional mean absolute deviation of 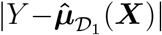 given the predictors, and 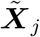 represents individual *j*’s covariates relevant to the error variance. Let 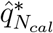 denote the ⌈(1 − *α*)(*N*_*cal*_ + 1)⌉-th smallest value among 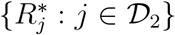. The corresponding prediction interval for a new target individual *N* + 1 is constructed as:

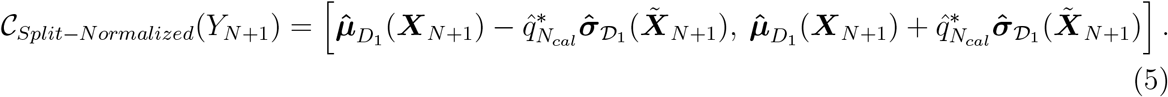

As demonstrated, the interval width is adaptive to the magnitude of the local error modeled by 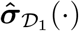. The critical challenge then lies in selecting an appropriate function to model the mean of the phenotypes and the heteroscedastic spread of residuals adequately.

CalPred developed a calibration model to construct context-specific prediction intervals^34^. It considers a calibration dataset ℐ and assumes that the phenotypes are normally distributed, with the conditional mean expressed as a linear combination of the predictors and the variance exponentially related to the relevant covariates. Under this framework, the phenotype is modeled as:

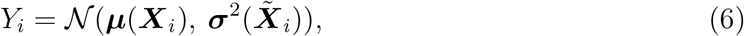

where 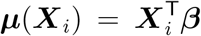 models the phenotypic mean using the predictors, and 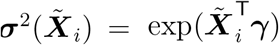 models the context-specific variance and the exponential function is used to ensure a non-negative variance. CalPred then estimates the parameters *β* and *γ* using the restricted maximum likelihood method to obtain the function estimators 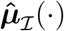 and 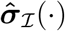 ^35^. Under the assumption of normality, the corresponding (1 − *α*)-level prediction interval is given by:

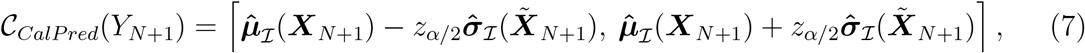

where *z*_*α/*2_ is the 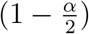-th quantile of a standard normal distribution.

Following the modeling approach of CalPred, we adopt a similar framework for phenotype prediction but relax the normality assumption. Specifically, we first fit a linear regression model to the phenotype *Y* using the predictors within the proper training set 𝒟_1_. With the fitted values, we can obtain the raw residuals. To model heteroscedasticity, we estimate the conditional variance by fitting a log-link Generalized Linear Model (GLM), regressing the squared residuals on relevant covariates. Subsequently, both fitted models are applied to the calibration set 𝒟_2_ to calculate the normalized residuals 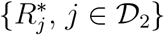. Finally, the estimated mean and variance functions are also applied to the target set 𝒟_*test*_, where prediction intervals are constructed by scaling the empirical residual quantile with the predicted local spread. The complete procedure of Split Conformal Prediction with Normalized Residuals (SCNR) is summarized in Algorithm 1.

#### Algorithm 1 Split Conformal Prediction with Normalized Residuals

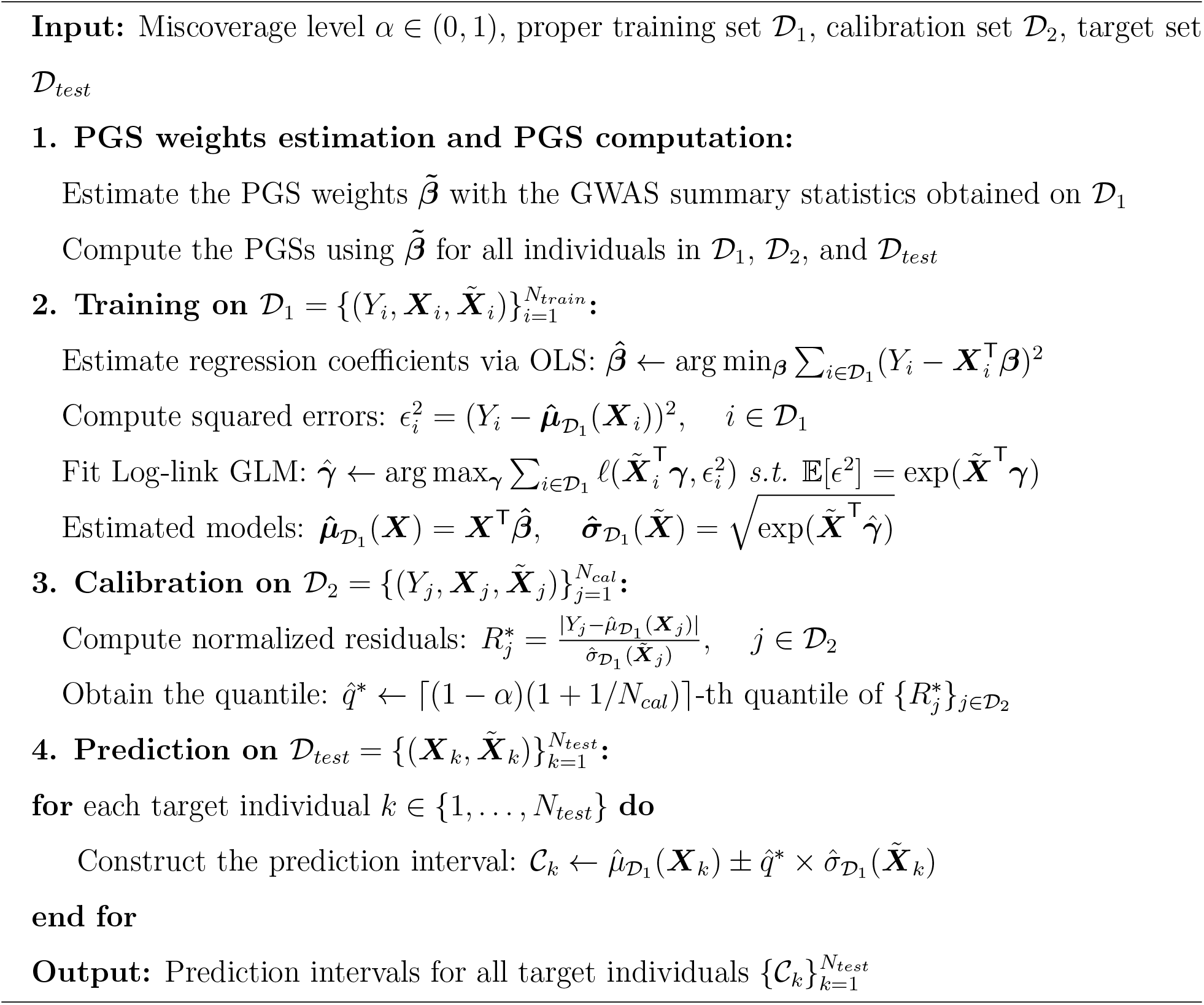

Despite the fact that Algorithm 1 can appropriately capture the broad heteroscedastic trends via GLM, the resulting prediction intervals are based on a global quantile obtained across the entire calibration set. Such an approach inherently assumes that once normalized, the residuals become exchangeable across the population, a premise that may falter if there exist latent subgroups with distinct error distributions that the mean and variance models fail to fully resolve. To address this problem, we introduce a stratified calibration strategy that partitions the data into more homogeneous clusters. A biologically intuitive implementation for such stratification is to define base groups according to key demographic interactions (e.g. Age × Sex). To ensure a data-driven and adaptive framework, we propose using Hierarchical Clustering to refine these strata. To avoid overfitting, the calibration set is first randomly partitioned into two equal-sized subsets, 𝒟_2,*A*_ and 𝒟_2,*B*_. The individuals in 𝒟_2,*A*_ are grouped into predefined base strata, and for each stratum, a corresponding feature vector is constructed using the normalized residuals. Here, we select two key features, the median and the tail-to-median ratio, to represent the local error profile. Next, Hierarchical Clustering is applied to these features to determine the optimal number of clusters. Clustering is performed using the R function *NbCluster()* with the Ward’s method and the Silhouette index. Subsequently, the learned cluster mapping is projected onto 𝒟_2,*B*_, where the cluster-specific quantiles are calculated. To enhance robustness and mitigate stochastic noise introduced by random splitting, we employ an ensemble-averaging technique. The ‘split-and-cluster’ process is repeated *L* times and the cluster-specific quantiles for all iterations are recorded. Finally, for each target individual, we identify the assigned cluster in each iteration and average the cluster-specific quantiles over the iterations. The ensemble quantile is then applied to construct the prediction interval. The pseudo-code of Clustering-based Split Conformal Prediction with Normalized Residuals (C-SCNR) is shown in Algorithm 2.

#### Algorithm 2 Clustering-based Split Conformal Prediction with Normalized Residuals

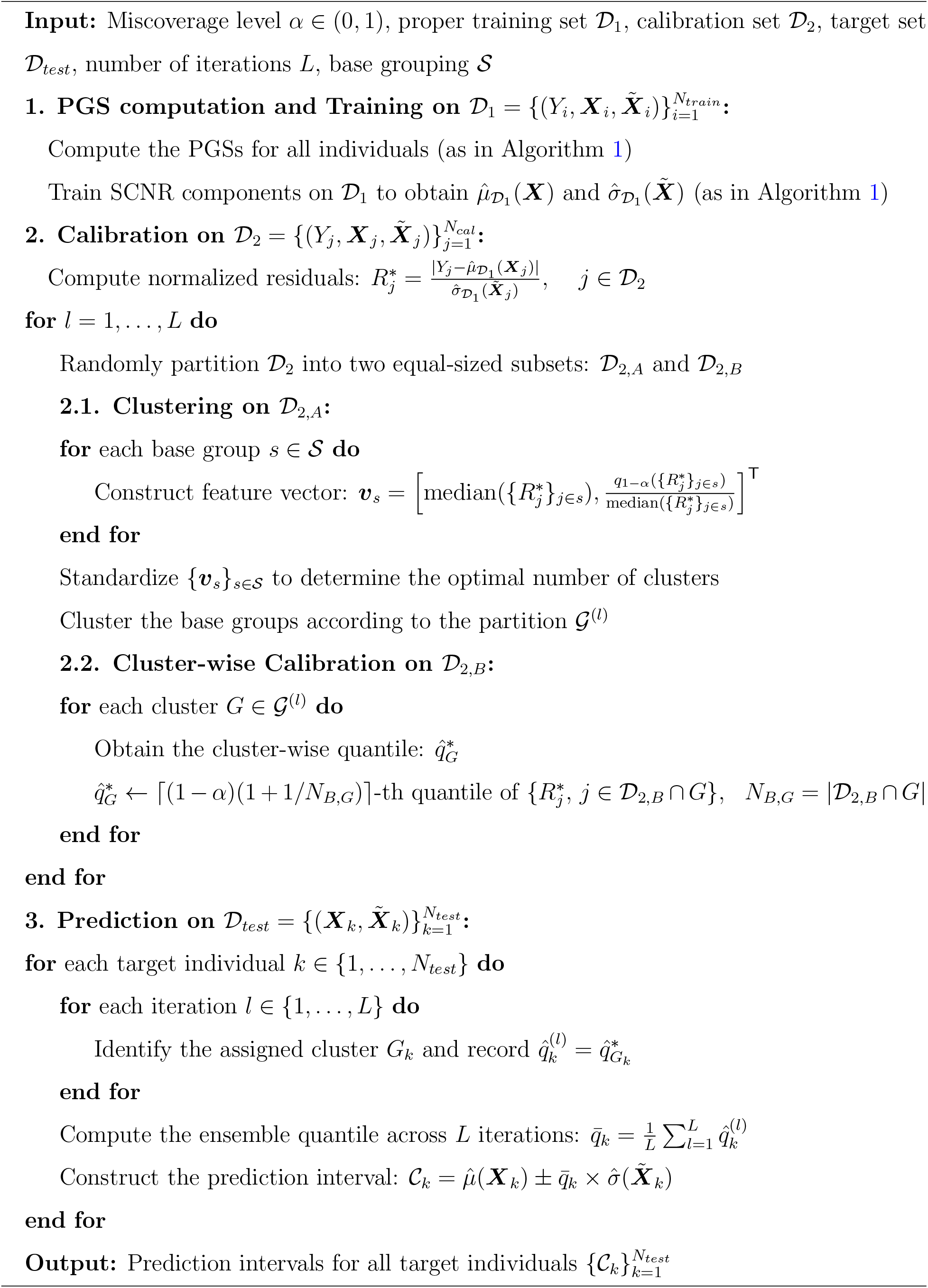

### UK Biobank Data Preprocessing

We followed the quality control (QC) protocols established by Li et al. ^38^ and Xu et al.^33^ to preocess the UK Biobank data. For sample QC, we retained individuals who (1) have available genotype data (data field 22005); (2) possess European ancestry (data field 21000). We excluded individuals who (1) have inconsistent genetic sex (data field 22001) and self-reported sex (data field 31); (2) are recommended to be excluded from genomic analysis (data field 22010); (3) are identified as outliers for heterozygosity or missing rate (data field 22027); (4) have close family relationships (data field 22018). For variant QC, we excluded variants with (1) a minor allele frequency (MAF) < 0.05; (2) an INFO score < 0.8; (3) a Hardy-Weinberg equilibrium (HWE) test *P* < 10^−6^; (4) a missing percentage > 5%; (5) duplicated SNPs; (6) multiple alleles. Moreover, we restricted our analysis to the intersection of the filtered SNPs and the HapMap3 SNPs^40^. After these QC procedures, a total of 314, 673 individuals and 1, 049, 629 SNPs remained for subsequent analyses. All the preprocessing steps were performed using PLINK2 (v2.00a6LM) ^39^.

### Compared Methods

We evaluated the performance of our proposed SCNR and C-SCNR frameworks against Split Conformal Prediction and existing state-of-the-art methods: PredInterval ^33^ and CalPred^34^. All the evaluated methods can be paired with any given PGS method. To ensure a fair and rigorous comparison, we employed a unified PGS estimation pipeline across all methods, utilizing the same PGS method, lassosum^29^, using the R package *lassosum* with default settings. The GWAS summary statistics for the training data were calculated using PLINK2 (v2.00a6LM)^39^ with `plink2 --glm`. The same reference panel was utilized to compute the Linkage Disequilibrium (LD) information. After estimating the PGS weights via lassosum, the PGS predictors in both the training and test data were obtained using PLINK (v1.90b7.2)^39^ with `plink --score`. For CalPred, we followed Hou et al. ^34^ and fitted CalPred on the same calibration set using the same set of covariates as in the Split Conformal Prediction-based methods. The quantile normalization for robust implementation was also applied, as suggested by Hou et al.^34^, to ensure the optimal performance for CalPred. For PredInterval, we followed Xu et al. ^33^ and implemented a 5-fold cross-validation. When predicting the phenotypic value, both for the training and test data, the set of predictors we used for PredInterval was kept identical to that employed for estimating the mean model in all other methods.

### Simulation Design

We conducted extensive simulation studies to evaluate the performance of our proposed methods in comparison with existing benchmarks. First, we randomly sampled *n* = 30, 000 individuals from the filtered UK Biobank data. We selected the SNPs on Chromosome 1 for these individuals with *m* = 85, 909 SNPs in total. We partitioned the 30, 000 individuals into a training set of 25, 000 individuals and a test set of 5, 000 individuals. The training set was further partitioned into 5 disjoint subsets of 5, 000 individuals for the 5-fold Cross-Validation, while we fixed one subset as the calibration set and treated the remaining four subsets as the proper training set for the Split Conformal Prediction framework. We randomly selected 1, 000 individuals from the remaining UK Biobank data to serve as a reference panel for the LD information. We obtained sex and age from the individual-level data accordingly. The PCs were computed on the selected panel of *n* individuals and *m* SNPs using PLINK2 (v2.00a6LM)^39^ via the command `plink2 --pca`. Next, we simulated quantitative phenotypes under various settings, including different genetic architectures, with or without covariate effects, and distinct error structures.

#### Simulation Study I: Comparison between Split Conformal Prediction and CV+

Simulation study I evaluates the performance of Split Conformal Prediction versus CV+ under standard settings. We followed the simulation design proposed by Xu et al. ^33^. The quantitative phenotype ***Y*** was generated using a standard linear polygenic model:

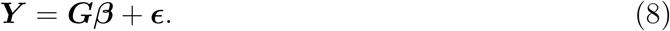

Here, ***G*** ∈ ℝ^*n*×*m*^ denotes the genotype matrix of *n* individuals across *m* SNPs. ***β*** = (*β*_1_,…,*β*_*m*_)^T^ represents the vector of effect sizes and ***ϵ*** = (*ϵ*_1_,…, *ϵ*_*n*_)^T^ denotes the residual error vector.

To simulate different genetic architectures, we introduced a polygenicity parameter *ρ* ∈ [0, 1], which dictates the proportion of causal variants. We first randomly selected a subset 𝒮_*causal*_ of *m*_*causal*_ = ⌊*mρ*⌋ SNPs to have non-zero effects. For these causal SNPs, their effect sizes are sampled independently from a normal distribution: 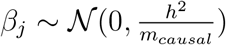, *j* ∈ 𝒮_*causal*_, where *h*^2^ denotes the heritability. For non-causal SNPs *j* ∉ 𝒮_*causal*_, we set *β*_*j*_ = 0. Subsequently, the effect sizes were back-transformed to the raw allelic scale by dividing each *β*_*j*_ by the expected genotype standard deviation 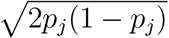, where *p*_*j*_ denotes the minor allele frequency (MAF) of SNP *j*. To maintain a total phenotypic variance of approximately 1, the residual errors were simulated from another normal distribution *ϵ*_*i*_ ∼ 𝒩 (0, 1 − *h*^2^), *i* ∈ [1,…, *n*]. Finally, we summed the genetic effects and residual errors to obtain the quantitative phenotypes.

To provide a comprehensive assessment, we first fixed the heritability at *h*^2^ = 0.5 and varied the polygenicity *ρ* ∈ {0.01, 0.1, 0.5} to examine the performance under varying degrees of genetic sparsity. Moreover, we fixed the polygenicity at *ρ* = 0.01 and varied the heritability *h*^2^ ∈ {0.2, 0.5, 0.8}, aiming to investigate the performance across different signal-to-noise ratios. We conducted ten simulation replicates for each setting.

#### Simulation Study II: Evaluation of Prediction Performance under Covariate Effects

Simulation study II investigates the prediction performance of SCNR, CalPred, and Split Conformal Prediction in the presence of covariate effects. We followed the design established by Hou et al.^34^ and Xu et al.^33^. The quantitative phenotype ***Y*** was simulated by incorporating covariates including sex, age, and PC1 from the UK Biobank data:

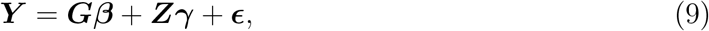

where ***Z*** is a covariate matrix comprising sex, age, and PC1, and ***γ*** denotes the vector of their effect sizes. Each column of ***Z*** was scaled to have zero mean and unit variance. To control the variance contribution, each covariate effect size was sampled from a normal distribution 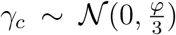, where *φ* signifies the proportion of phenotypic variance attributed to the covariates. A central aspect of this simulation study is the evaluation of method robustness against non-constant residual variance. Therefore, we simulated the residual errors from a heteroscedastic normal distribution where the variance is a function of the covariates. The variance of individual *i* is modeled as: 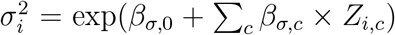. The residual errors were sampled from 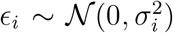, *i* ∈ [1,…,*n*]. Following the parameterization proposed by Hou et al.^34^, we set the coefficients to *β*_*σ*,sex_ = 0.2, *β*_*σ*,age_ = 0.25, and *β*_*σ*,PC1_ = 0.15. The baseline intercept *β*_*σ*,0_ was selected to ensure that the aggregate residual variance equals to 1 −*h*^2^ −*φ*. We varied the variance proportion *φ* ∈ {0.05, 0.1, 0.15} to simulate different levels of variance contributed by these covariates. Furthermore, the heritability and polygenicity were also varied across *h*^2^ ∈ {0.2, 0.5, 0.8} and *ρ* ∈ {0.01, 0.1, 0.5}. For each parameter configuration, we performed ten independent simulation replicates and assessed the context-specific prediction coverage rate and average interval width across these replicates.

#### Simulation Study III: Evaluation of Robustness to Skewed Errors

Simulation Study III explores the performance of SCNR and CalPred in scenarios where the normality assumption is violated. Following the design established by Hou et al.^34^, we first simulated a point estimate *Ŷ*_*i*_ ∼ 𝒩 (0, 1) for each individual *i*. We utilized the real context from the UKB data including sex, age, and PC1, which were scaled to have mean 0 and variance 1. To introduce skewness into the phenotype, instead of a normal distribution, we simulated the errors using a centered and scaled exponential distribution. For each individual *i*, the phenotype was generated as:

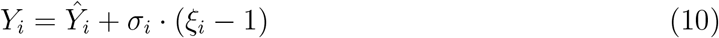

where *ξ*_*i*_ is sampled from an exponential distribution with a rate parameter of 1, i.e., *ξ*_*i*_ ∼ Exp(1). This construction ensures that the residual has a zero mean and a context-specific variance of 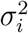, which is modeled in the same manner as described in Simulation Study The baseline intercept *β*_*σ*,0_ was selected such that *R*^2^(***Y***, ***Ŷ***) = 0.3 for individuals with average covariate values (∑_*c*_ *β*_*σ,c*_ × *Z*_*i,c*_ = 0). The simulation was replicated twenty times, and we evaluated the performance of SCNR and CalPred under different confidence levels. We varied the target coverage levels from 0.2 to 0.99 (1 − *α* ∈ {0.2, 0.4, 0.6, 0.8, 0.9, 0.99}), following Xu et al.^33^.

#### Simulation Study IV: Validation of C-SCNR

Simulation Study IV aims to illustrate the effectiveness of C-SCNR through group-level comparisons with SCNR. We designed a series of simulation scenarios featuring stratified error structures. The phenotype *Y*_*i*_ for each individual *i* was generated as:

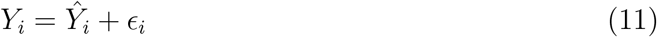

where *Ŷ*_*i*_ ∼ 𝒩 (0, 1) and the residual term *ϵ*_*i*_ is sampled from another normal distribution 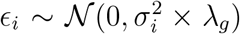. In this framework, 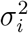 represents the baseline heteroscedastic variance driven by covariates, which is modeled as in the previous simulation studies, while *λ*_*g*_ denotes a group-specific scaling factor.

In Scenario 1, we implemented a 2×2 factorial design representing the interaction between sex (S0: female; S1: male) and age (A1: age < 55; A2: age ≥ 55), resulting in four distinct base groups. The scaling factor was set to *λ*_*g*_ = 0.5 for younger groups (S0A1, S1A1) and *λ*_*g*_ = 2 for older groups (S0A2, S1A2). Scenario 2 extended this to a 2 × 3 design by discretizing age into three tiers (A1: age < 50; A2: age 50 − 60; A3: age *>* 60), yielding six base groups where the scaling factor *λ*_*g*_ was set as 0.5 for A1, 1.0 for A2, and 2.0 for A3. Finally, Scenario 3 adopted a 2 × 2 × 2 design by incorporating genetic ancestry via a binary split of PC1 at the median (P1: PC1 < median; P2: PC1 ≥ median), creating eight base groups from S0A1P1 to S1A2P2. Similarly, *λ*_*g*_ ∈ {0.5, 1.0, 2.0} were assigned to simulate three heterogeneous error strata. The simulation was replicated twenty times for each scenario, and the number of iterations was set to *L* = 20 for C-SCNR. The performances of C-SCNR and SCNR were evaluated across the predefined base groups.

Furthermore, we conducted a sensitivity analysis to investigate the impact of different numbers of ‘split-and-cluster’ iterations. We generated the phenotype according to procedure in Simulation Study II, using normally distributed context-specific residuals. Parameters were fixed at *h*^2^ = 0.5, *ρ* = 0.01, and *φ* = 0.1. We implemented a 2 × 6 factorial design based on the interaction between sex (S0: female; S1: male) and age (A1: age < 45; A2: age 45 − 50; A3: age 50 − 55; A4: age 55 − 60; A5: age 60 − 65; A6: age > 65). We varied the number of iterations *L* ∈ {10, 20, 50}, and the simulation was replicated ten times.

### Real Data Application

We analyzed three quantitative traits from the UK Biobank to evaluate the performance of our proposed methods and the existing methods. We selected a physical measurement (BMI) and two lipid traits (LDL and HDL). The overview of the analyzed phenotypes is summarized in the Supplementary Table. Following Xu et al. ^33^, we first randomly sampled 1, 000 individuals from the preprocessed UK Biobank data to serve as a reference panel for computing the LD information. For each trait, individuals with missing values were removed. Subsequently, we partitioned the remaining individuals into five disjoint subsets of equal size. Each subset served as the test set in turn, resulting in five independent trials for each trait. Consistent with our simulation design, the remaining data is treated as the training data and is further divided into five equally sized folds to implement a 5-fold CV. For C-SCNR and SCNR, one of the five folds was designated as the calibration set and the remaining four folds constituted the proper training set. CalPred was applied using the same calibration set.

We obtained the GWAS summary statistics using PLINK2 (v2.00a6LM) ^39^ with the command plink2 `--glm`, adjusting for sex, age, and the top 20 PCs as covariates. The top PCs were computed on the preprocessed panel using `plink2 --pca`. For all evaluated methods, the PGS weights were obtained using lassosum^29^ with default settings, and the PGSs for both the training and test datasets were calculated using PLINK (v1.90b7.2)^39^ with `plink --score`. For model fitting, we followed Hou et al.^34^, estimated the mean function using PGS, age, age × sex, age^2^, and the top 10 PCs as predictors. The variance function was fitted with predictors include age, sex, PC1 and PC2. To evaluate localized performance, we defined twelve base groups through a 2 × 6 interaction between sex (S0: female; S1: male) and age (A1: age < 45; A2: age 45 − 50; A3: age 50 − 55; A4: age 55 − 60; A5: age 60 − 65; A6: age > 65), where the age splits were driven by the distribution of UKB data. When performing hierarchical clustering in C-SCNR, we set the minimum and maximum number of clusters to 1 and 4, ensuring flexibility while maintaining computational stability. For BMI and LDL, we primarily benchmarked C-SCNR against SCNR. For HDL particularly, we conducted an exhaustive comparison involving C-SCNR, SCNR, CalPred, PredInterval, and a modified version of PredInterval incorporating normalized residuals.

## 3 Results

### Simulation Results

#### Simulation Study I

The results of Simulation Study I indicate that the Split Conformal Prediction and CV+ (implemented via PredInterval) achieve comparable performance across a wide range of simulated genetic architectures (Figure 1). As demonstrated in the left panels of Figure 1A and 1B, both frameworks consistently maintain an empirical coverage precisely at the target 0.95 level. The robustness is consistently observed across varying levels of polygenicity and heritability. Regarding efficiency, the right panels of Figure 1A and 1B show that the mean interval widths are virtually identical between the compared frameworks. While Split Conformal Prediction exhibits a slightly larger spread in interval widths under some settings, their median values remain essentially equivalent.

**Figure 1.**
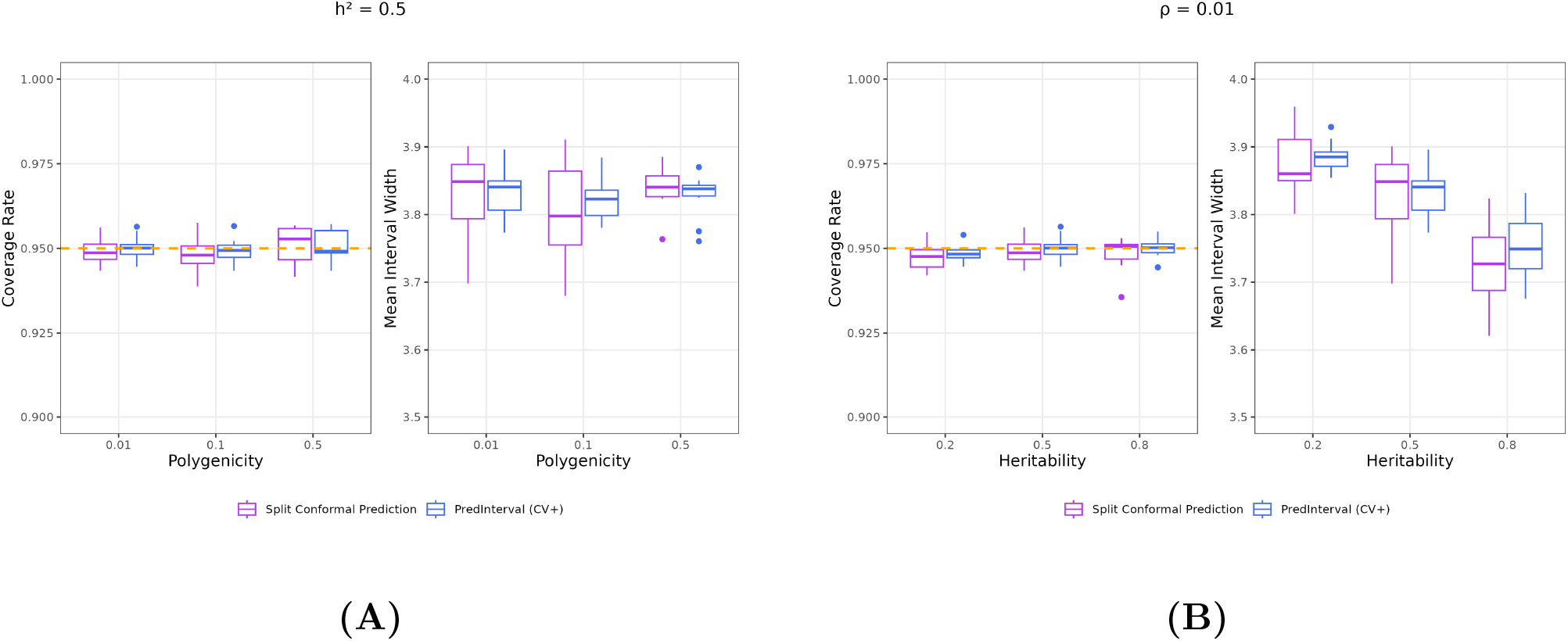
Performance comparison between Split Conformal Prediction and CV+. Simulations were performed with a training set of 25, 000 individuals. The prediction intervals for a target set of 5, 000 individuals were evaluated. The box plots were constructed based on 10 simulation replicates for each setting, where the center line indicates the median and the box limits represent the 25% and 75% percentiles. The dashed orange lines represent the target 0.95 coverage level. Each panel displays the empirical prediction coverage rate (left) and the corresponding mean interval width (right). **(A)** Varying polygenicity values (*ρ*) = 0.01, 0.1, 0.5 with heritability (*h*^2^) fixed at 0.5; **(B)** Varying heritability (*h*^2^) = 0.2, 0.5, 0.8 with polygenicity (*ρ*) set to 0.01.

Under the simulated settings considered here, CV+ and Split Conformal Prediction show nearly identical performance. Therefore, for computational simplicity, we use Split Conformal Prediction as the main global conformal baseline in the subsequent simulation studies.

#### Simulation Study II

The results of Simulation Study II demonstrate that evaluating prediction performance solely at the population level can mask significant disparities in conditional validity in the presence of heteroscedasticity (Figure 2 and Supplementary Figures 1-4). As shown in Figure 2A, SCNR, CalPred, and Split Conformal Prediction all achieve the target 0.95 coverage rate when averaged across the entire cohort, and the mean interval widths present minor discrepancies. However, a more granular analysis reveals that this overall-level success for Split Conformal Prediction is achieved through a trade-off that results in over-coverage in certain subgroups and under-coverage in others.

**Figure 2.**
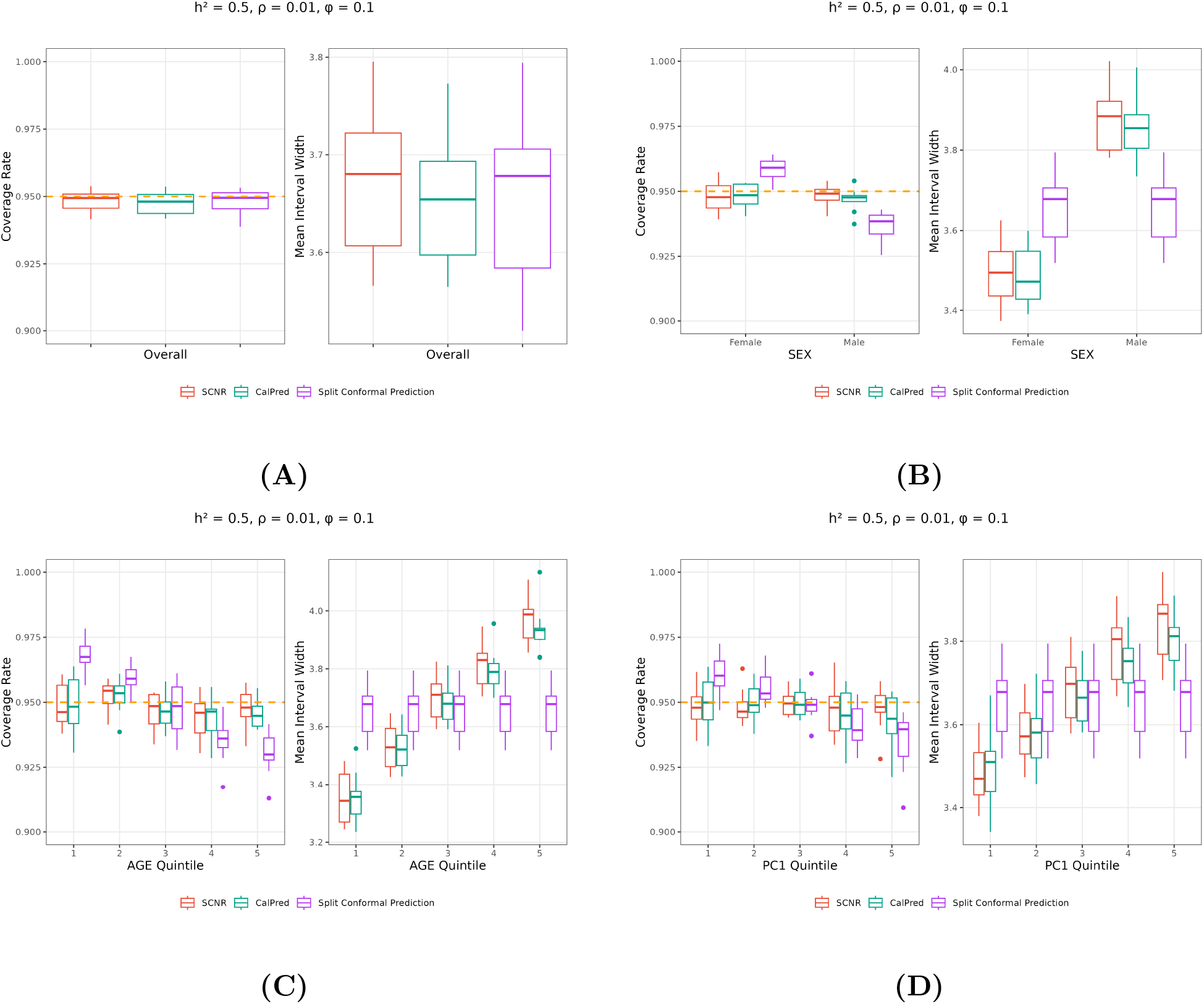
Performance comparison between SCNR, CalPred, and Split Conformal Prediction. Simulations were performed with a proper training set of 20, 000 individuals and a calibration set of 5, 000 individuals. The prediction intervals for 5, 000 testing individuals were evaluated for each context subgroup (quintiles of age and PC1; female/male for sex). The box plots were constructed based on the 10 simulation replicates for each setting. The dashed orange lines represent the target 0.95 confidence level. The parameters are set to heritability *h*^2^ = 0.5, polygenicity *ρ* = 0.01, and proportion of variance attributed to covariates *φ* = 0.1. Each panel displays the empirical prediction coverage rate (left) and the corresponding mean interval width (right). **(A)** At the overall level; **(B)** subgroups stratified by sex; **(C)** subgroups stratified by quintiles of age; **(D)** subgroups stratified by quintiles of PC1.

Specifically, Split Conformal Prediction exhibits unstable coverage when the data is displayed through covariate stratification (Figure 2B, 2C, and 2D). For instance, the intervals provided by Split Conformal Prediction are overly conservative for females, resulting in excessive coverage, whereas those for males are too narrow to meet the target level. This trend persists across age and PC1 quintiles, as Split Conformal Prediction fails to provide valid intervals for subgroups with higher residual variance. Consistent with the introduced theories, Split Conformal Prediction tends to provide intervals of constant width across subgroups, as a result of not accounting for conditional variance. In contrast, both SCNR and CalPred maintain a stable coverage rate of 0.95 across all subgroups, demonstrating superior local adaptivity. Notably, since *φ* modulates the contribution of covariates to the mean rather than the residual variance, varying its value introduces negligible shifts to the coverage behaviors (Figure 2, Supplementary Figures 1 and 2). Instead, the heritability *h*^2^ acts as the primary driver of the substantial variations observed across the parameter configurations, particularly dictating the extent of localized miscalibration for Split Conformal Prediction (Supplementary Figures 3 and 4). Crucially, SCNR and CalPred consistently anchor the coverage rate at the target level throughout these diverse genetic architectures.

This disparity highlights the fundamental tension between interval efficiency and validity. While narrower intervals are generally preferred, our results suggest that efficiency should not be prioritized at the expense of validity. For higher-variance subgroups, SCNR and CalPred provide systematically wider intervals. Rather than representing a loss of efficiency, such adjustments are necessary to maintain valid coverage under heteroscedastic conditions. Meanwhile, the narrower intervals constructed for lower-variance subgroups reflect an adaptive adjustment to local data distributions, and the higher interval efficiency is more of a byproduct. By dynamically scaling intervals based on the specific context of each individual, our proposed framework ensures that the prediction intervals remain robust and reliable for all participants, whereas the constant-width intervals might be misleadingly precise or poorly calibrated.

#### Simulation Study III

The results of Simulation Study III are presented in Figure 3. Under a skewed error distribution, SCNR demonstrates remarkable stability across a range of target coverage levels. For both male and female subgroups, the empirical coverage rates of SCNR adhere closely to the reference line. This consistency indicates that SCNR is capable of accommodating non-normal error structures.

**Figure 3.**
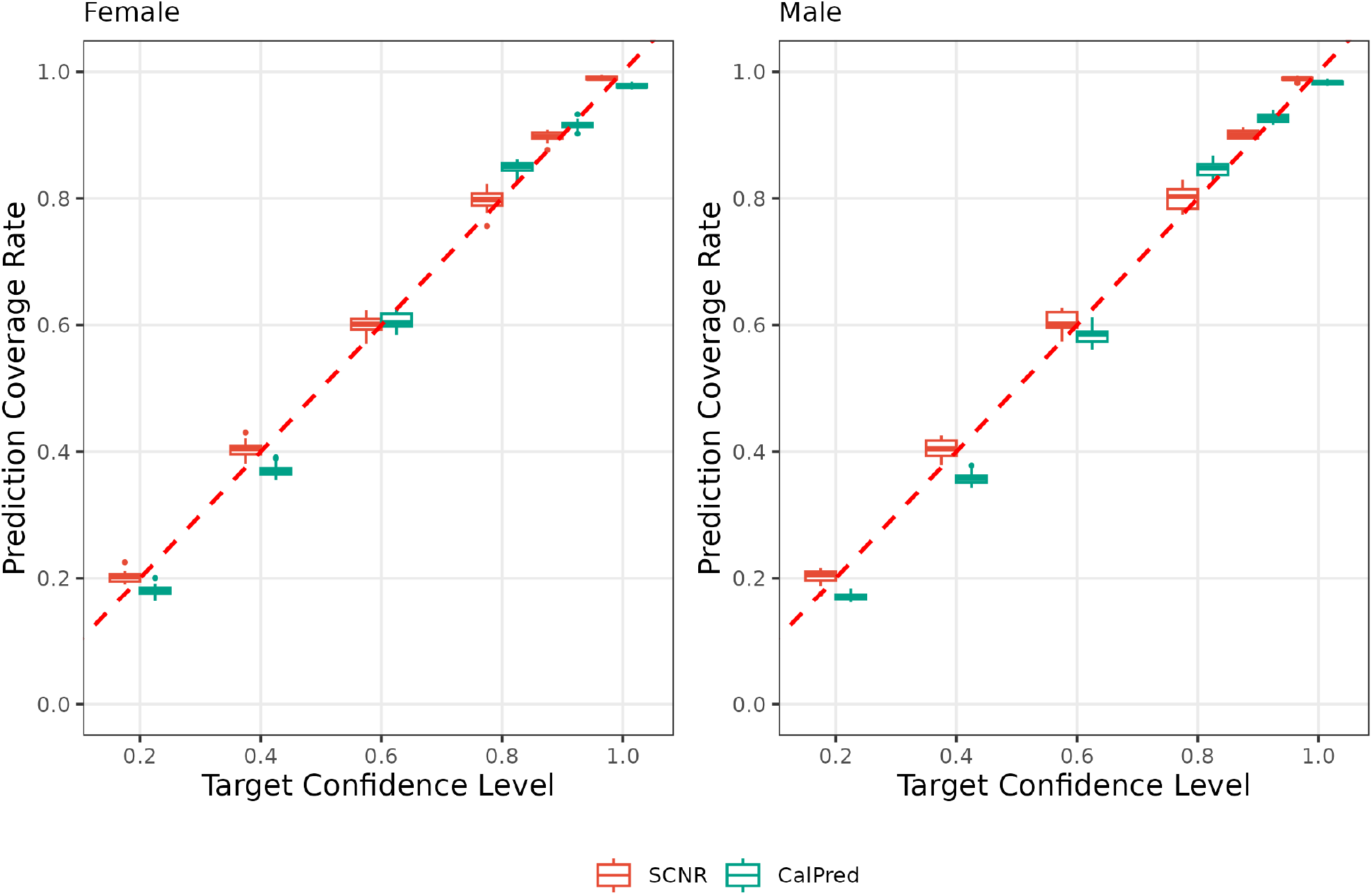
Performance comparison between SCNR and CalPred under skewed error distributions. Simulations were performed with a proper training set of 20, 000 individuals and a calibration set of 5, 000 individuals. The prediction intervals for 5, 000 testing individuals were evaluated for context subgroups. Box plots were constructed based on 20 simulation replicates. The dashed red line refers to empirical coverage rate matching the target level. Each panel displays empirical prediction coverage rate versus different values of target confidence level. **(Left)** Female; **(Right)** Male.

In contrast, although implemented with quantile normalization, CalPred exhibits instability in coverage across the considered levels. At lower target levels (≤ 0.6), CalPred systematically under-covers the phenotype. Conversely, it tends to over-cover at higher target levels (0.8, 0.9). These systematic deviations suggest that while addressing heteroscedasticity, CalPred remains sensitive to the underlying distribution and its accuracy relies on normality. The adaptive approach of SCNR offers a more robust framework for uncertainty quantification, ensuring that the prediction intervals remain valid regardless of the target confidence level and the presence of underlying skewness.

#### Simulation Study IV

As shown in Figure 4, C-SCNR demonstrates superior adaptive calibration to stratified error structures compared to SCNR across all three scenarios. In the simplest case (Figure 4A), SCNR over-covers low-variance groups and fails to reach the target coverage level for high-variance groups. In contrast, C-SCNR provides valid prediction intervals for all base groups. For more complex cases (Figure 4B and 4C), although not achieving perfect calibration, C-SCNR consistently adjusts the intervals for each base group, bringing the empirical coverage closer to the target. Since SCNR applies a global quantile after modeling contexts into the residual variance, it cannot resolve the localized stratification simulated here. However, C-SCNR effectively identifies the latent strata via the ‘split-and-cluster’ procedure and dynamically scales the intervals based on the clustering results.

**Figure 4.**
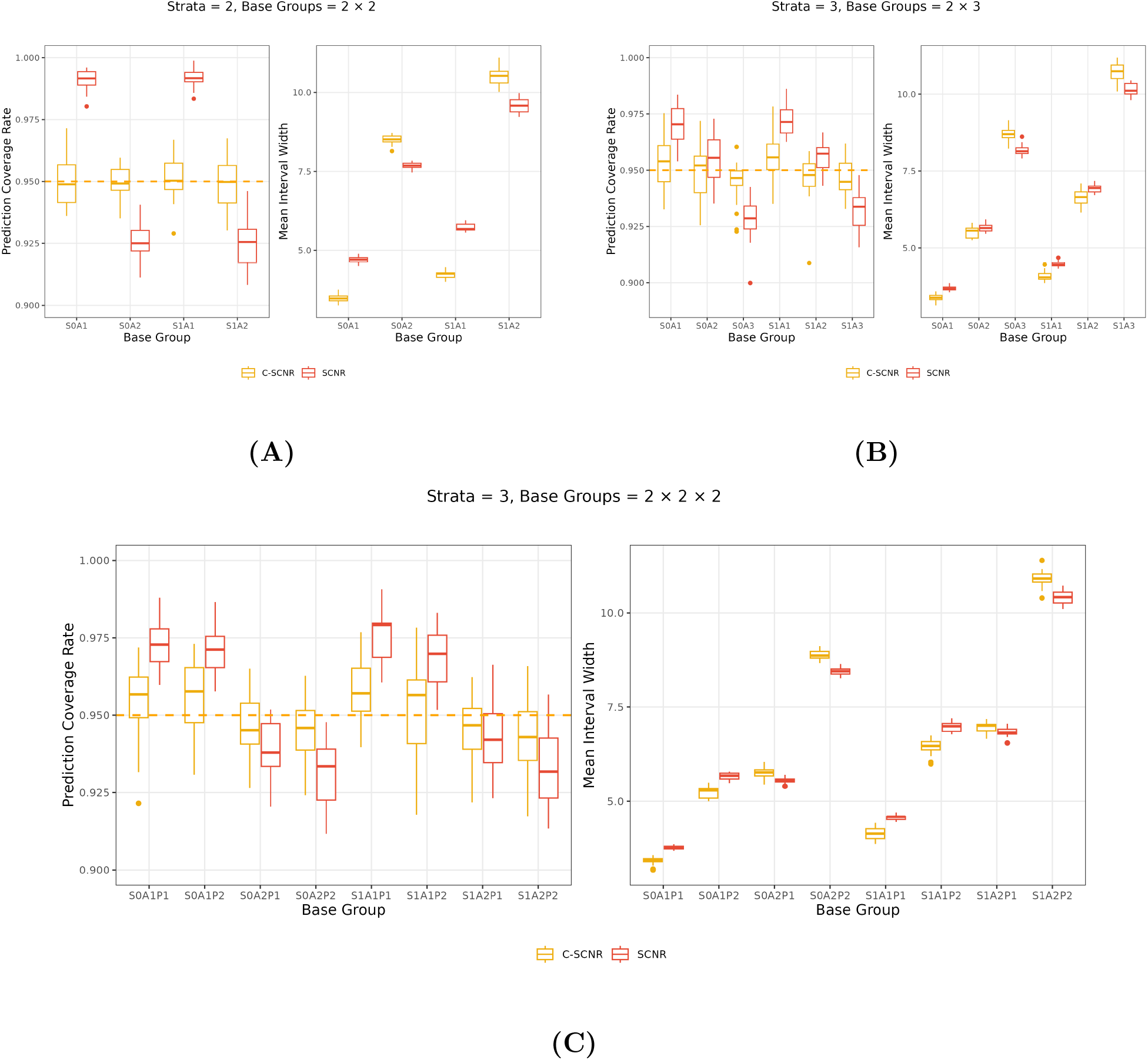
Performance comparison between C-SCNR and SCNR under stratified error structures. Simulations were performed with a proper training set of 20, 000 individuals and a calibration set of 5, 000 individuals. The prediction intervals for 5, 000 testing individuals were evaluated for input base groups. Box plots were constructed based on 20 simulation replicates for each configuration. The dashed orange lines represent the target 0.95 confidence level. Each panel displays the empirical prediction coverage rate (left) and the corresponding mean interval width (right) by predefined base groups. **(A)** Sex × Age (2 × 2) design with 2 error strata; **(B)** Sex × Age (2 × 3) design with 3 error strata; **(B)** Sex × Age × PC1 (2 × 2 × 2) design with 3 error strata.

The trade-off between localized validity and efficiency is reflected in the interval widths (Figure 4A, 4B, and 4C). For the groups that SCNR under-covers, C-SCNR appropriately constructs wider intervals to capture the true phenotypic dispersion. Conversely, in the lower-variance tiers where SCNR is unnecessarily conservative, C-SCNR yields narrower and more precise intervals. This pattern remains robust as the complexity of the design increases, and even in the most challenging scenario (Figure 4C), C-SCNR significantly improves the coverage rate towards the target 0.95 level. These results validate C-SCNR as a powerful extension of the Split Conformal Prediction framework, capable of providing reliable uncertainty quantification even when the underlying structure is characterized by stratified, covariate-driven discontinuities.

The results of the sensitivity analysis are shown in Figure 5. As illustrated, the empirical coverage rates and mean interval widths remain nearly invariant as *L* ranges from 10 to 50. In real data applications, we recommend using *L* = 20 for a balance between stability and computational efficiency. Moreover, Figure 5 also demonstrates the performance of C-SCNR in the absence of a specifically designed error structure (i.e., a standard heteroscedastic scenario). C-SCNR consistently maintains the nominal 0.95 coverage rate across all twelve base groups. This suggests that C-SCNR inherits the advantages of SCNR and serves ad a versatile framework that can produce reliable intervals across diverse heteroscedastic landscapes, regardless of whether the residual variance is normal, skewed, or stratified.

**Figure 5.**
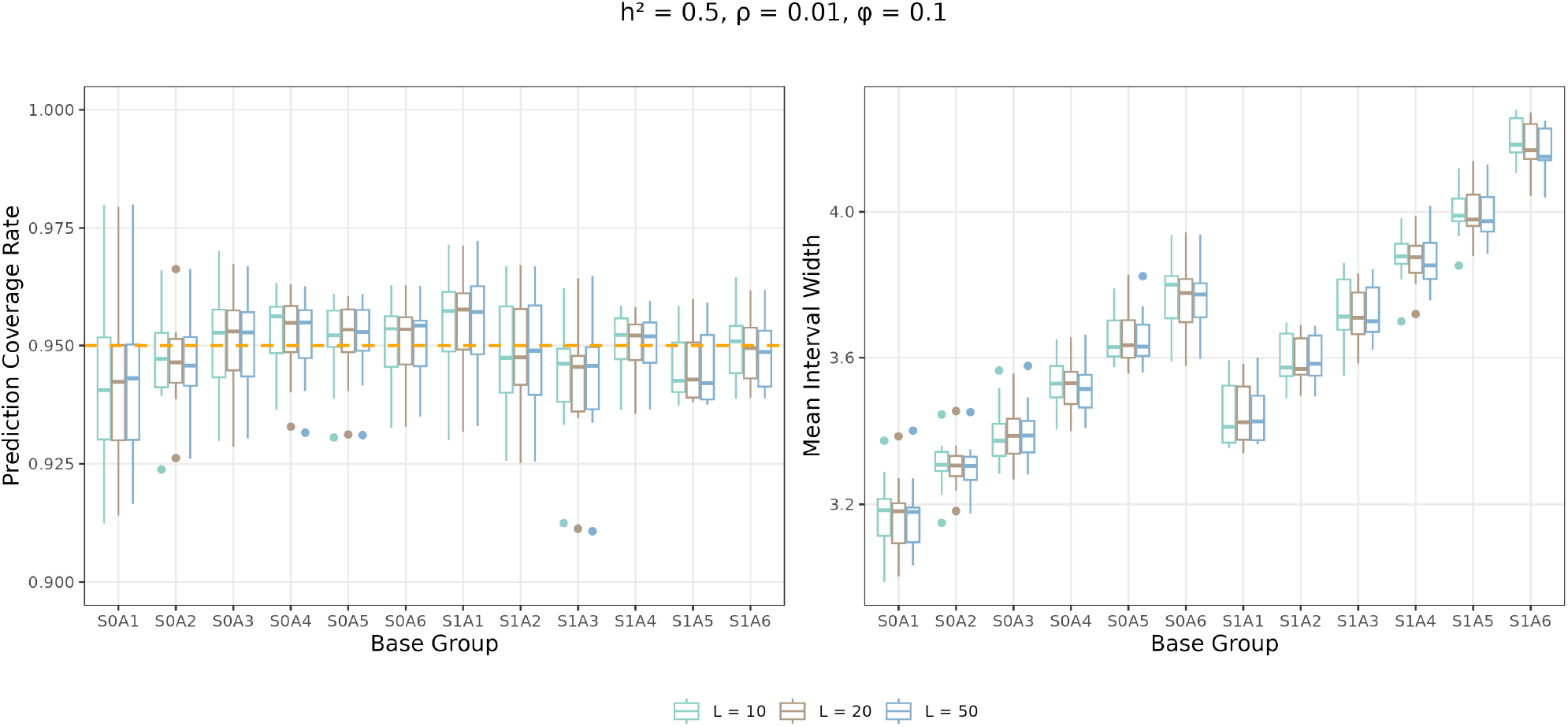
Sensitivity analysis of the number of split-and-cluster iterations (*L*) on C-SCNR performance. Simulations were performed with a proper training set of 20, 000 individuals and a calibration set of 5, 000 individuals. The prediction intervals for 5, 000 testing individuals were evaluated for input base groups. Box plots were constructed based on 10 simulation replicates. The dashed orange lines represent the target 0.95 confidence level. The heritability, polygenicity, and proportion of variance attributed to covariates were set to *h*^2^ = 0.5, *ρ* = 0.01, and *φ* = 0.1. Each panel displays the performance metrics across different numbers of repetitions *L* ∈ {10, 20, 50}. **(Left)** Empirical prediction coverage rate; **(Right)** Mean interval width.

### Real Data Results

The prediction performances of C-SCNR and SCNR for the UK Biobank traits BMI and LDL are presented in Figure 6 and 7. The results indicate that the C-SCNR framework offers consistent advantages over SCNR in real data applications. For BMI, C-SCNR provides wider prediction intervals for groups such as S0A3, S0A4, S1A3, and S1A4, which leads to coverage rates that align more closely with the target 0.95 level (Figure 6A and 6B). These results highlight the importance of identifying localized stratification for individuals aged between 50 and 60. Meanwhile, the intervals yielded by C-SCNR are significantly narrower for the S0A6, S1A1, and S1A2 groups, bringing the coverage rates closer to the 0.95 target. For most remaining groups, C-SCNR yields coverage rates that track the 0.95 target level more accurately, although no significant differences in the mean widths are observed. The advantages are more pronounced for LDL. Specifically, the narrower intervals produced for groups including S0A1, S0A2, and S1A6 demonstrate that C-SCNR avoids overly conservative coverage while remaining valid (Figure 7A and 7B). Furthermore, wider intervals adjust the coverage rates upward towards 0.95, even for the S1A1 group that is the hardest to calibrate. While SCNR serves as a strong baseline by accounting for heteroscedasticity, it exhibits localized calibration gaps that C-SCNR has the capacity to resolve. These findings are highly consistent with our observations in Simulation Study IV, validating that the C-SCNR framework is better equipped to handle the latent variance stratification inherent in complex quantitative traits.

**Figure 6.**
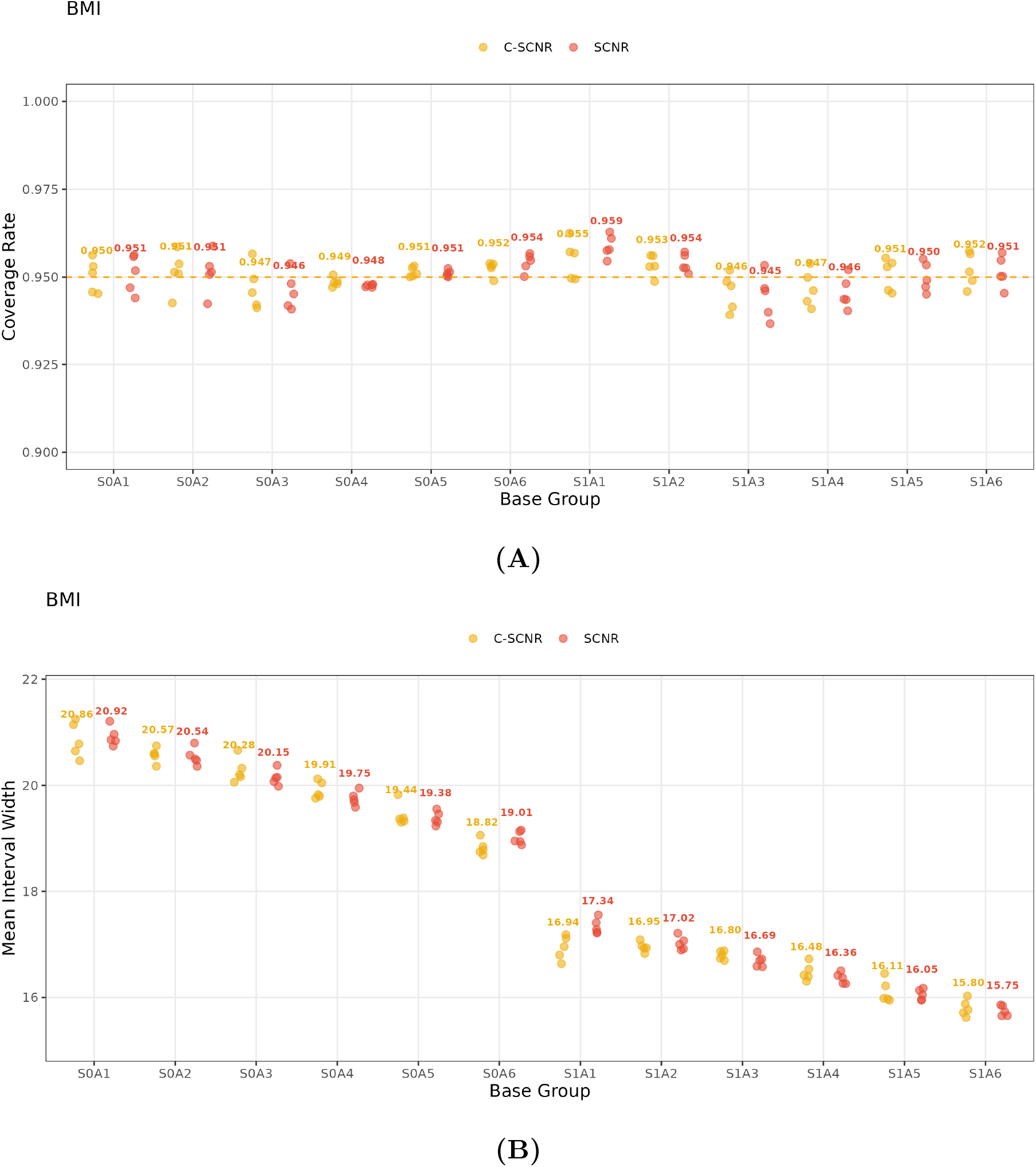
Performance comparison between C-SCNR and SCNR for Body mass index (BMI). The jitter plot displays the prediction performance for five independent trials using UK Biobank individual-level data across the 12 predefined base groups. Mean values for each group are displayed above the points. **(A)** Empirical prediction coverage rate; **(B)** Mean interval width.

**Figure 7.**
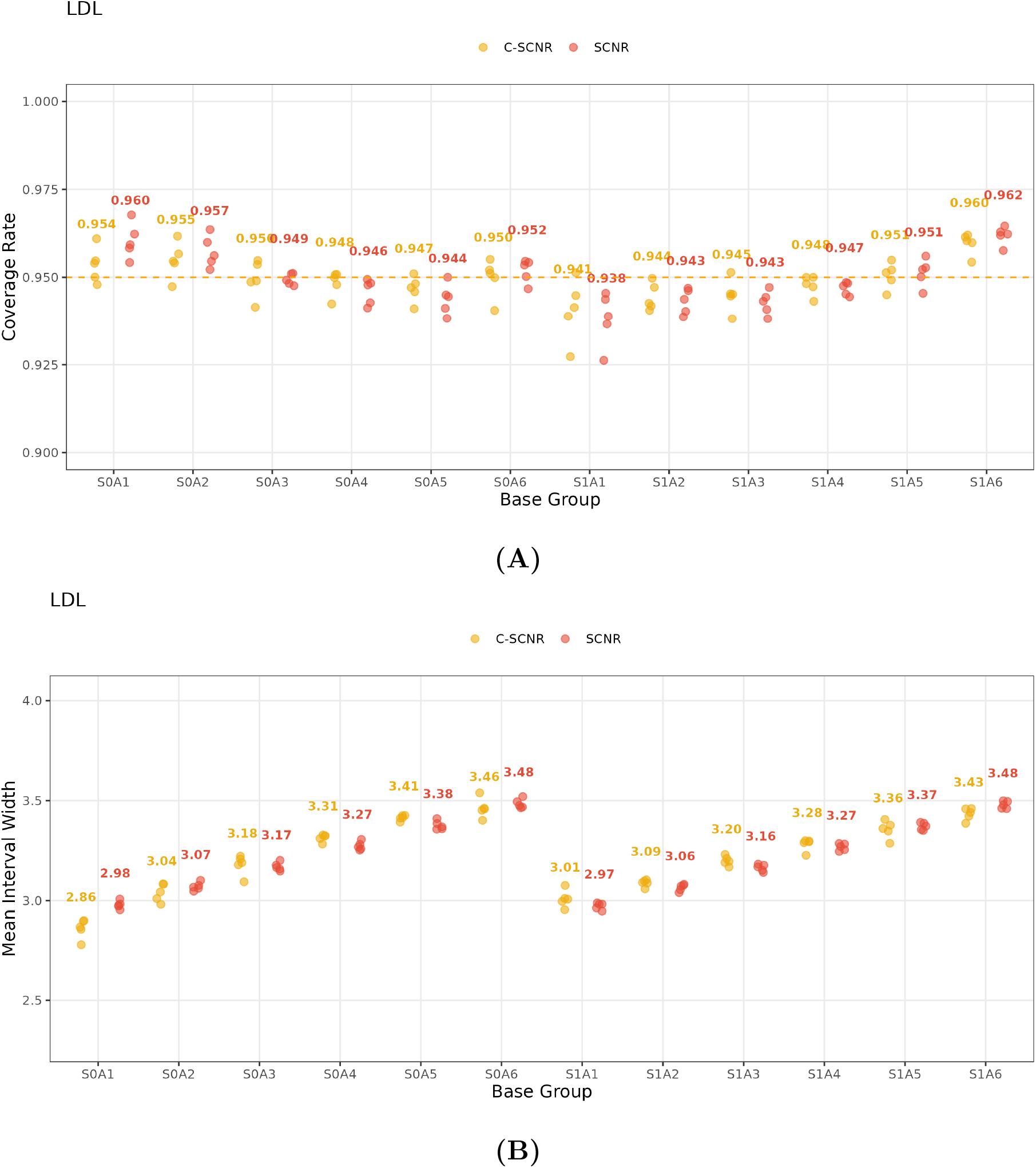
Performance comparison between C-SCNR and SCNR for Low-density lipoprotein (LDL) cholesterol. The jitter plot displays the prediction performance for five independent trials using UK Biobank individual-level data across the 12 predefined base groups. Mean values for each group are displayed above the points. **(A)** Empirical prediction coverage rate; **(B)** Mean interval width.

The comprehensive evaluation of HDL highlights the varying capabilities of the existing frameworks and our proposed methods (Figure 8). PredInterval exhibits suboptimal performance with coverage rates fluctuating sharply from 0.926 to 0.976 across different base groups. This is directly attributable to its roughly static interval widths across demographic strata, which fail to adapt to the underlying heteroscedasticity (Figure 8B). When incorporating normalized residuals (PredInterval+NR), results improve markedly, becoming comparable to those of SCNR. C-SCNR, SCNR, and CalPred all demonstrate high localized validity, maintaining coverage near the 0.95 target (Figure 8A). Notably, C-SCNR provides a marginal but consistent improvement over SCNR and CalPred, particularly in the male base groups. This performance does not indicate a limitation of C-SCNR, but rather underscores the architectural flexibility of its framework. In scenarios where the stratification effects are subtle, the ‘split-and-cluster’ procedure does not introduce bias or instability to the prediction intervals. Instead, it captures nuanced local variations or naturally defaults to fewer clusters, still offering refined interval calibration without compromising robustness.

**Figure 8.**
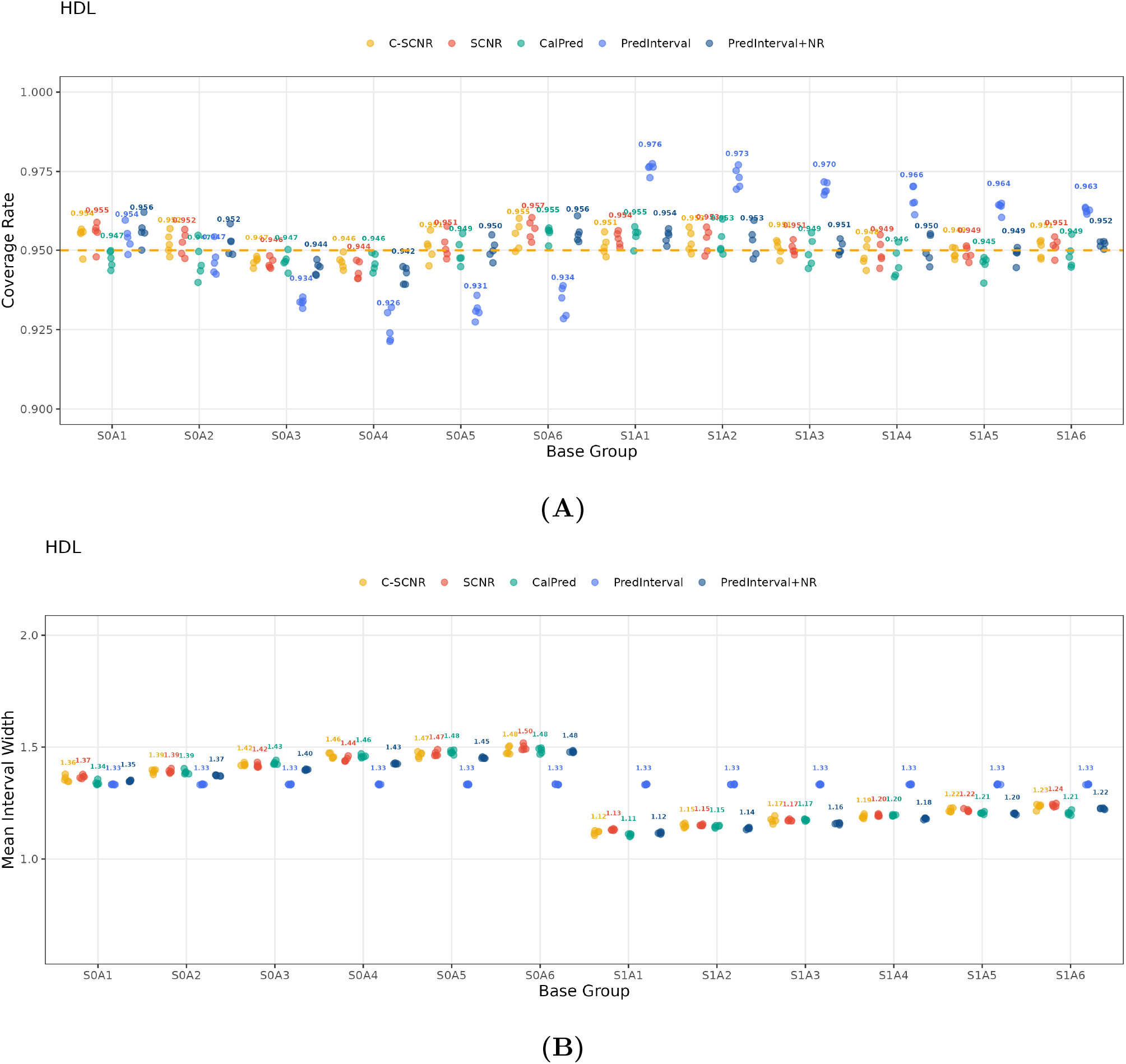
Performance comparison of different methods for High-density lipoprotein (HDL) cholesterol. The jitter plot displays the prediction performance for five independent trials using UK Biobank individual-level data across the 12 predefined base groups. Mean values for each group are displayed above the points. **(A)** Empirical prediction coverage rate; **(B)** Mean interval width.

## 4 Discussion

In this study, we have introduced C-SCNR, a framework designed to construct locally-calibrated phenotype prediction intervals. Building upon the Split Conformal Prediction framework, our approach addresses the challenge of heteroscedasticity in phenotypic prediction. We first proposed SCNR, which utilizes an estimated variance function to normalize residuals, adequately accounting for broad heteroscedasticity driven by covariates such as age, sex, and genetic ancestry. In Simulation Study II, we demonstrated the effectiveness of SCNR via the incorporation of normalized residuals. This modeling approach outperformed the standard Split Conformal Prediction and CV+ framework, which were shown as equivalent in Simulation Study I. Furthermore, Simulation Study III verified that SCNR is more robust to non-normal phenotypic distributions than CalPred. Recognizing that stratified variance is prevalent in complex biological traits and that global normalization alone cannot fully capture such structures, we extended to a clustering-based framework, C-SCNR. By incorporating a ‘split-and-cluster’ mechanism and aggregating the results across multiple random partitions, C-SCNR achieves a more granular level of calibration, rendering it robust to covariate-driven discontinuities in the error distributions. Simulation Study IV revealed that while SCNR may suffer from localized calibration gaps in the presence of stratified error structures, C-SCNR effectively resolves these gaps by dynamically identifying the underlying latent strata.

The real data application results validated our simulation findings, as C-SCNR demonstrated consistent advantages over SCNR and other methods across the predefined base groups. For BMI and LDL, C-SCNR mitigated the under-coverage and over-coverage issues within specific demographic strata, most notably for individuals aged between 50 and 60 for BMI and females under fifty for LDL. Crucially, the HDL analysis showed that in scenarios where variance shifts are relatively smooth, C-SCNR avoided performance degradation and maintained stable prediction performance comparable to SCNR and CalPred across all base groups. This success is attributable to the flexibility of our framework. C-SCNR accepts customized base groups as input, which can be tailored for specific phenotypes based on biological insights. Our framework also enables data-driven self-learning of stratification. Such locally-adaptive prediction intervals are pivotal for clinical decision-making, as they ensure validity for individuals across detailed demographic backgrounds rather than merely at the population macro-level.

Despite these advantages, several limitations warrant consideration. First, the implementation of our framework has a higher demand on data availability than existing methods. Unlike PredInterval and CalPred, which can operate with summary-level statistics or small calibrations sets, our framework requires the individual-level data to accurately model the variance function for residual normalization. While individual-level data are essential for high-resolution localized calibration, this requirement may limit the method’s applicability in settings where only summary statistics are available. Second, the analysis of HDL suggests that the predefined base groups constructed solely by sex and age interactions may not fully capture the heterogeneity inherent in all phenotypes. Primary sources of heteroscedasticity may also stem from other factors, such as lifestyle, environmental exposures and medication usage, where higher-order covariate interactions should be considered. Nevertheless, C-SCNR allows for the flexible inclusion of additional covariates into the base group definitions, ensuring that this framework can be refined to achieve more precise, biologically-informed calibration. Third, our work focuses on quantitative traits only and extending this framework to binary outcomes remains a non-trivial challenge. Among the existing approaches, CalPred attempts to model the liability for disease traits. While theoretically sound, it complicates the direct interpretation of predictions in clinical practice. Alternatively, PredInterval treats the binary outcomes as continuous, which ensures the nominal coverage but often results in excessively wide intervals that lack practical utility. We believe that developing a more rigorous and interpretable approach for binary trait prediction is a critical future extension. Finally, the current study focuses exclusively on individuals of European ancestry within the UK Biobank. Challenges arise when the data include individuals from different ancestral groups. Future extensions of our work should aim to adapt the C-SCNR framework to multi-ancestry settings, ensuring equitable and accurate uncertainty quantification across populations.

## Data Availability

All data produced in the present study are available upon reasonable request to the authors.

## Data availability

The UK Biobank data are accessed under Application Number 140822. The individual-level genotype and phenotype data are available at https://www.ukbiobank.ac.uk/. The HapMap3 SNP list is available at https://ftp.ncbi.nlm.nih.gov/hapmap/.

## Code availability

SCNR and C-SCNR are available at https://github.com/yunyurain/C-SCNR. CalPred is available at https://github.com/KangchengHou/calpred. PredInterval is available at https://github.com/xuchang0201/PredInterval. lassosum is available at https://github.com/tshmak/lassosum. PLINK (v1.9) is available at https://www.cog-genomics.org/plink/. PLINK (v2.0) is available at https://www.cog-genomics.org/plink/2.0/.

## Author contributions

Y.Y. and Y.D.Z. conceived the project. Y.Y. developed the methodology, performed the simulation studies and real data applications, and drafted the manuscript. Y.D.Z. critically revised the manuscript and supervised the work. X.H. provided critical feedback and revised the manuscript.

