## Supplementary Figures and Table for "Locally adaptive conformal prediction intervals for polygenic score-based phenotype prediction via residual normalization and data-driven stratification"

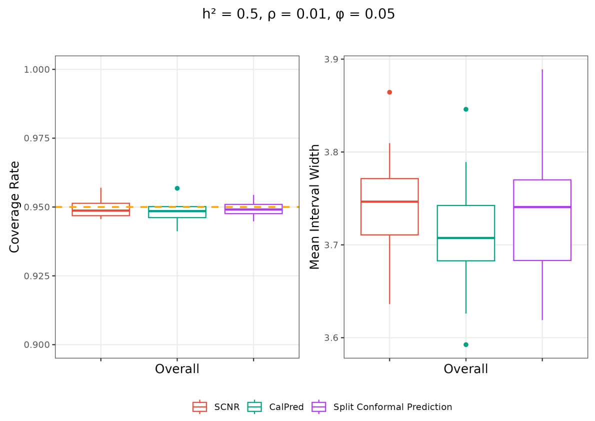


**(A)**

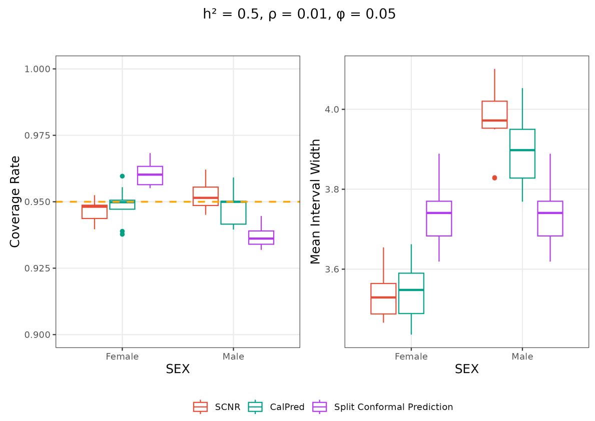


**(B)**

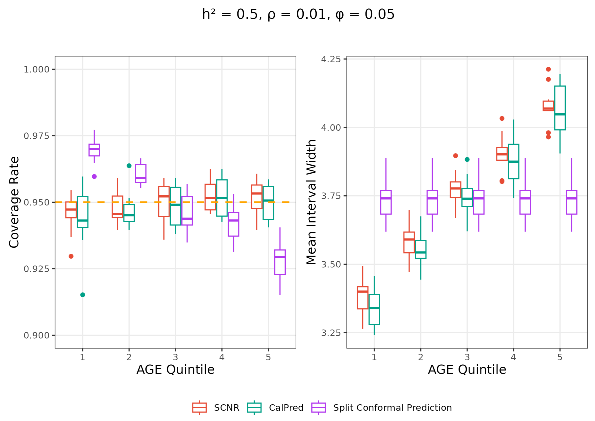


**(C)**

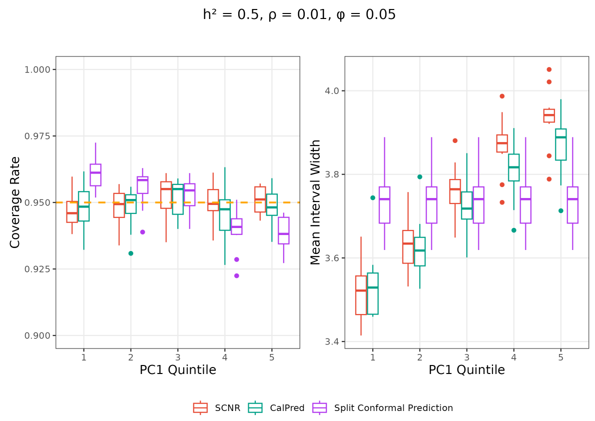


**(D)**

**Supplementary Figure 1. Performance comparison between SCNR, CalPred, and Split Conformal Prediction.** Simulations were performed with a proper training set of $20,000$ individuals and a calibration set of $5,000$ individuals. The prediction intervals for $5,000$ testing individuals were evaluated for each context subgroup (quintiles of age and PC1; female/male for sex). The box plots were constructed based on the 10 simulation replicates for each setting. The dashed orange lines represent the target 0.95 confidence level. The parameters are set to heritability $h^2 = 0.5$, polygenicity $\rho = 0.01$, and proportion of variance attributed to covariates $\varphi = 0.05$. Each panel displays the empirical prediction coverage rate (left) and the corresponding mean interval width (right). **(A)** At the overall level; **(B)** subgroups stratified by sex; **(C)** subgroups stratified by quintiles of age; **(D)** subgroups stratified by quintiles of PC1.

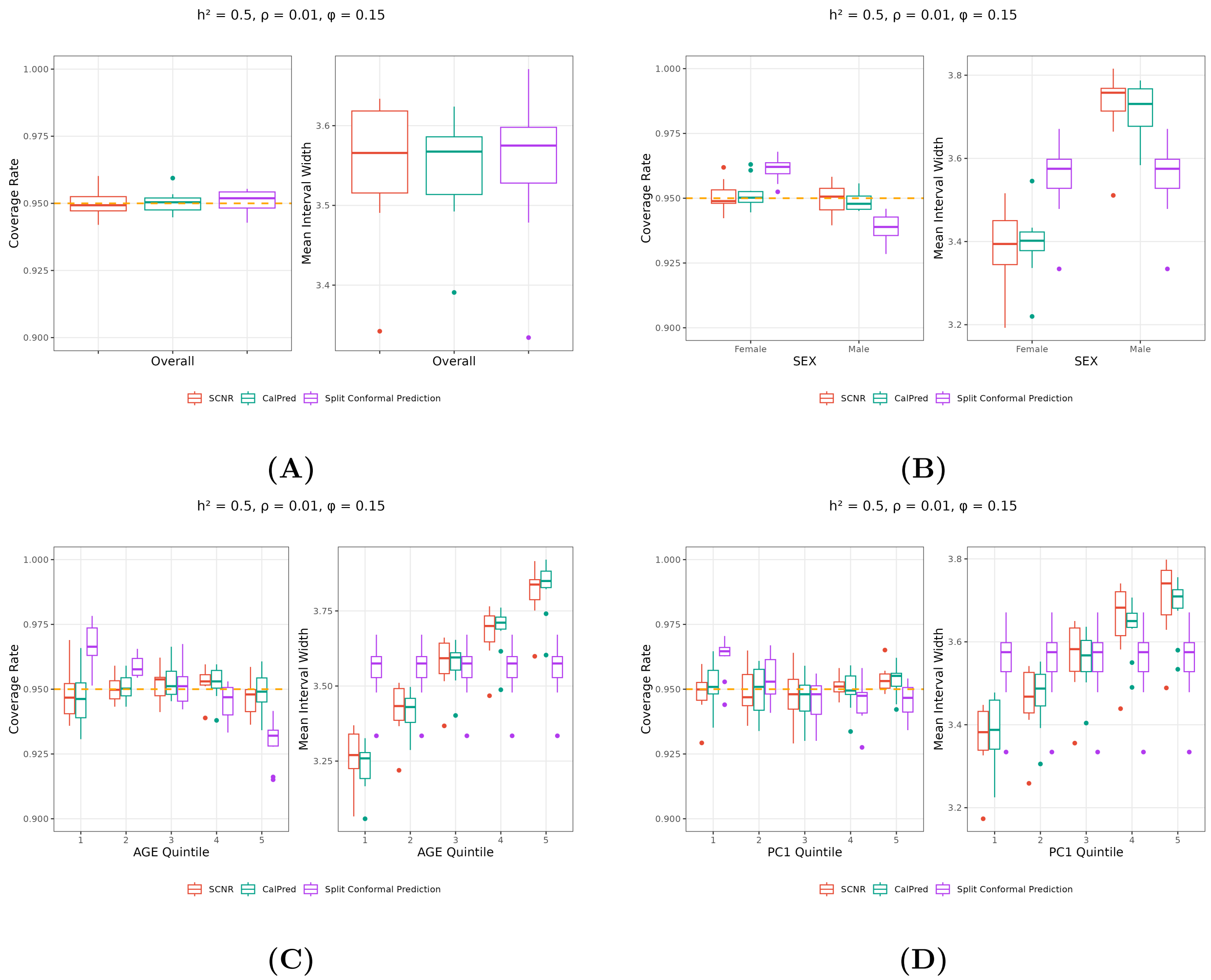


**Supplementary Figure 2. Performance comparison between SCNR, CalPred, and Split Conformal Prediction.** Simulations were performed with a proper training set of 20,000 individuals and a calibration set of 5,000 individuals. The prediction intervals for 5,000 testing individuals were evaluated for each context subgroup (quintiles of age and PC1; female/male for sex). The box plots were constructed based on the 10 simulation replicates for each setting. The dashed orange lines represent the target 0.95 confidence level. The parameters are set to heritability $h^2 = 0.5$, polygenicity $\rho = 0.01$, and proportion of variance attributed to covariates $\varphi = 0.15$. Each panel displays the empirical prediction coverage rate (left) and the corresponding mean interval width (right). **(A)** At the overall level; **(B)** subgroups stratified by sex; **(C)** subgroups stratified by quintiles of age; **(D)** subgroups stratified by quintiles of PC1.

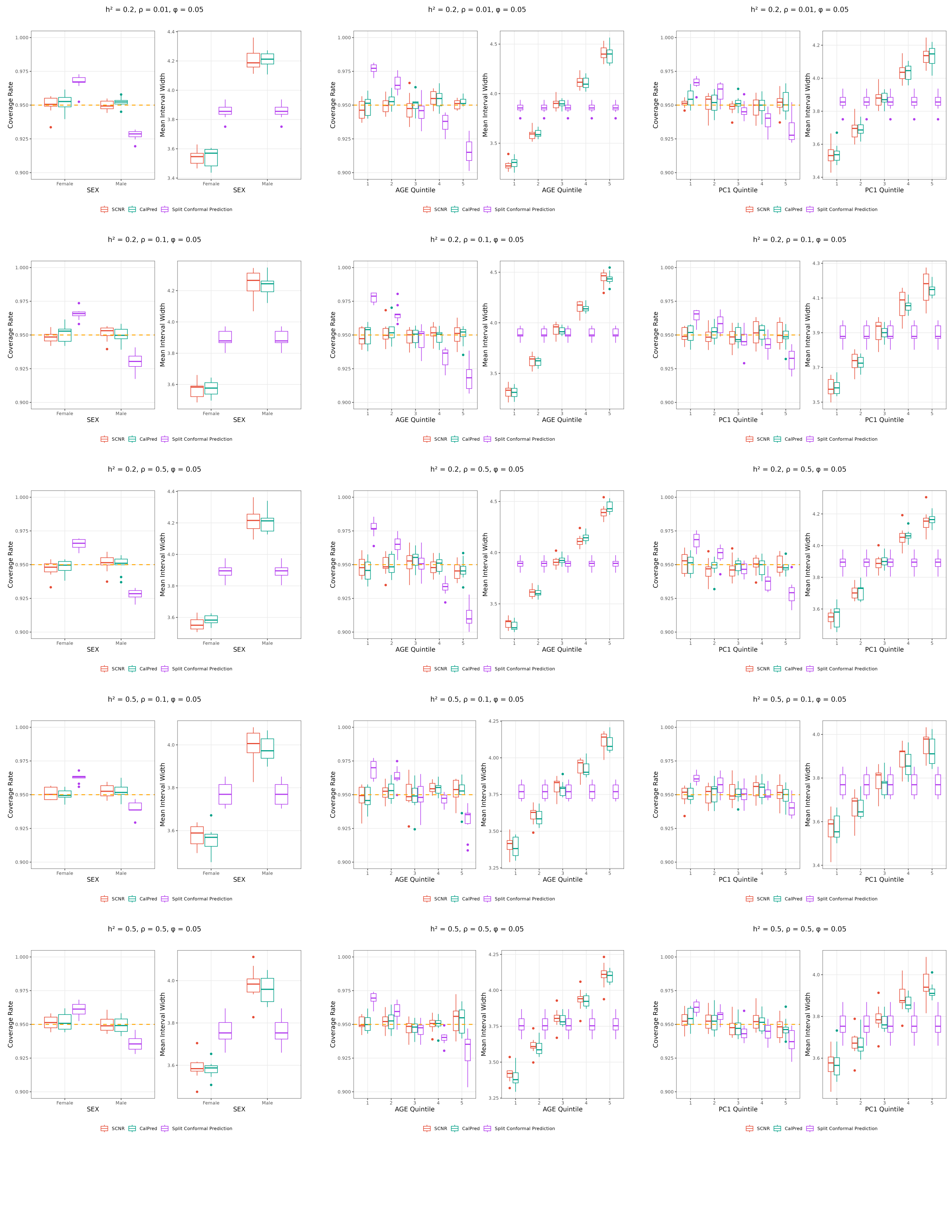

h² = 0.2, ρ = 0.01, φ = 0.05
h² = 0.2, ρ = 0.1, φ = 0.05
h² = 0.2, ρ = 0.5, φ = 0.05
h² = 0.5, ρ = 0.1, φ = 0.05
h² = 0.5, ρ = 0.5, φ = 0.05
Coverage Rate
Mean Interval Width
SEX
Female
Male
AGE Quintile
PC1 Quintile
SCNR
CalPred
Split Conformal Prediction

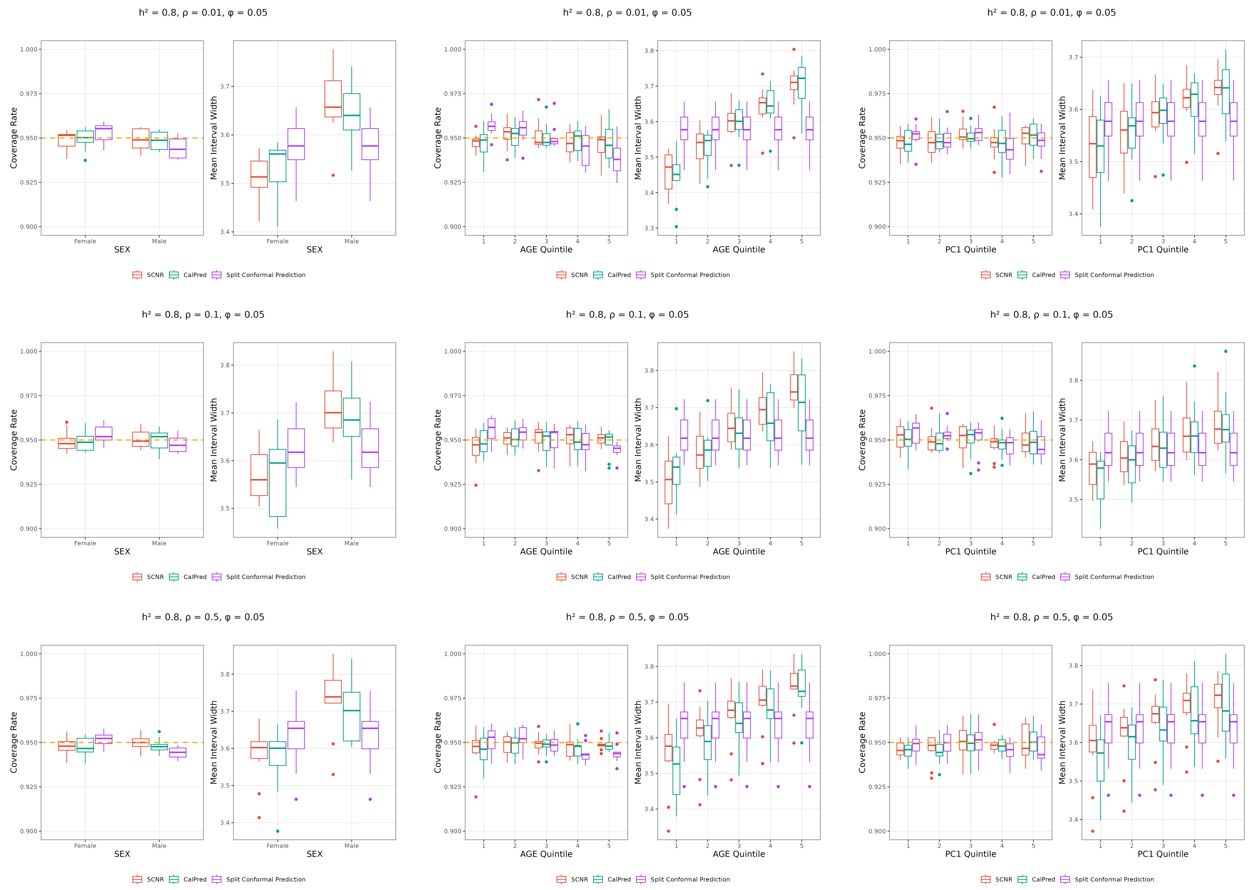


**Supplementary Figure 3. Performance comparison between SCNR, CalPred, and Split Conformal Prediction.** Simulations were performed with a proper training set of $20,000$ individuals and a calibration set of $5,000$ individuals. The prediction intervals for $5,000$ testing individuals were evaluated for each context subgroup (quintiles of age and PC1; female/male for sex). The box plots were constructed based on the 10 simulation replicates for each setting. The dashed orange lines represent the target 0.95 confidence level. The parameters are set to heritability $h^2 = \{0.2, 0.5, 0.8\}$, polygenicity $\rho = \{0.01, 0.1, 0.5\}$, and proportion of variance attributed to covariates $\varphi = 0.05$. Each panel displays the empirical prediction coverage rate (left) and the corresponding mean interval width (right). The panels in each row correspond to one parameter configuration, where **(Left)** subgroups stratified by sex; **(Middle)** subgroups stratified by quintiles of age; **(Right)** subgroups stratified by quintiles of PC1.

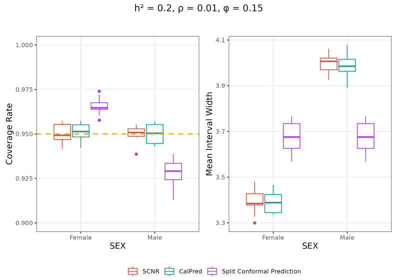
$h^2 = 0.2, \rho = 0.01, \varphi = 0.15$
Coverage Rate
Mean Interval Width
SEX
SCNR
CalPred
Split Conformal Prediction

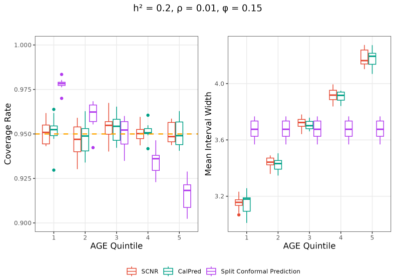
$h^2 = 0.2, \rho = 0.01, \varphi = 0.15$
Coverage Rate
Mean Interval Width
AGE Quintile
SCNR
CalPred
Split Conformal Prediction

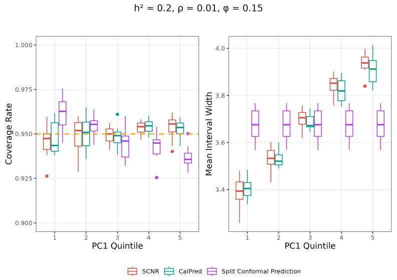
$h^2 = 0.2, \rho = 0.01, \varphi = 0.15$
Coverage Rate
Mean Interval Width
PC1 Quintile
SCNR
CalPred
Split Conformal Prediction

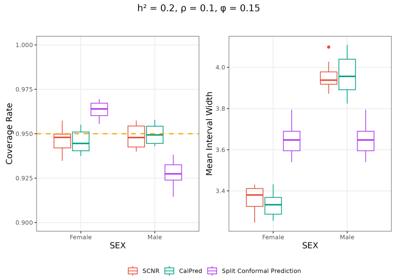
$h^2 = 0.2, \rho = 0.1, \varphi = 0.15$
Coverage Rate
Mean Interval Width
SEX
SCNR
CalPred
Split Conformal Prediction

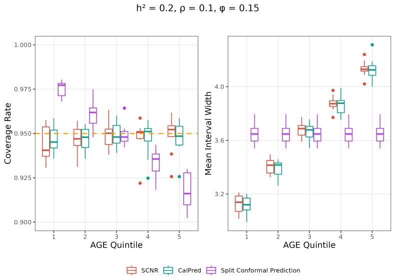
$h^2 = 0.2, \rho = 0.1, \varphi = 0.15$
Coverage Rate
Mean Interval Width
AGE Quintile
SCNR
CalPred
Split Conformal Prediction

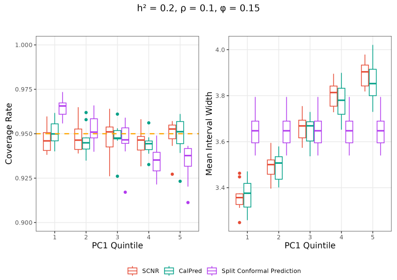
$h^2 = 0.2, \rho = 0.1, \varphi = 0.15$
Coverage Rate
Mean Interval Width
PC1 Quintile
SCNR
CalPred
Split Conformal Prediction

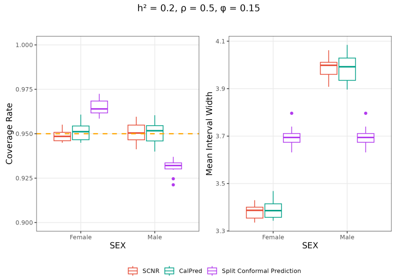
$h^2 = 0.2, \rho = 0.5, \varphi = 0.15$
Coverage Rate
Mean Interval Width
SEX
SCNR
CalPred
Split Conformal Prediction

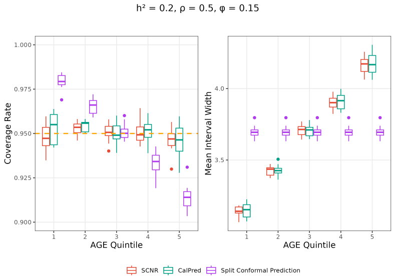
$h^2 = 0.2, \rho = 0.5, \varphi = 0.15$
Coverage Rate
Mean Interval Width
AGE Quintile
SCNR
CalPred
Split Conformal Prediction

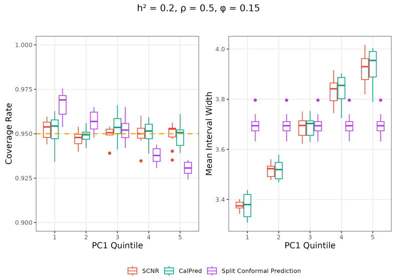
$h^2 = 0.2, \rho = 0.5, \varphi = 0.15$
Coverage Rate
Mean Interval Width
PC1 Quintile
SCNR
CalPred
Split Conformal Prediction

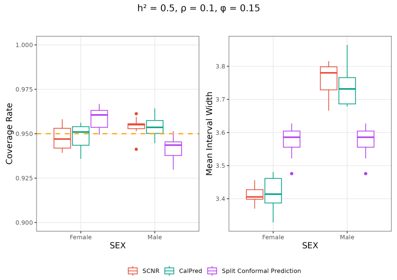
$h^2 = 0.5, \rho = 0.1, \varphi = 0.15$
Coverage Rate
Mean Interval Width
SEX
SCNR
CalPred
Split Conformal Prediction

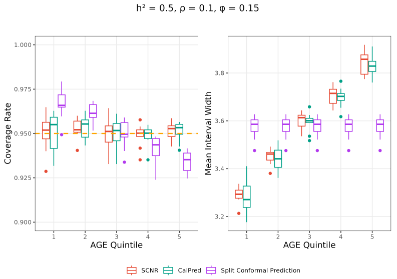
$h^2 = 0.5, \rho = 0.1, \varphi = 0.15$
Coverage Rate
Mean Interval Width
AGE Quintile
SCNR
CalPred
Split Conformal Prediction

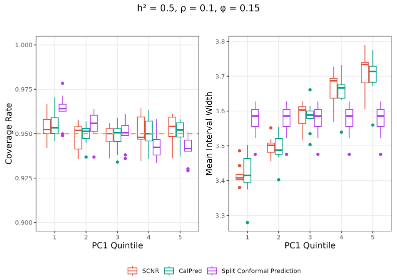
$h^2 = 0.5, \rho = 0.1, \varphi = 0.15$
Coverage Rate
Mean Interval Width
PC1 Quintile
SCNR
CalPred
Split Conformal Prediction

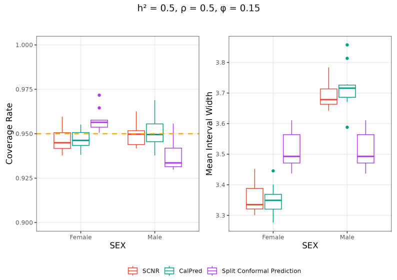
$h^2 = 0.5, \rho = 0.5, \varphi = 0.15$
Coverage Rate
Mean Interval Width
SEX
SCNR
CalPred
Split Conformal Prediction

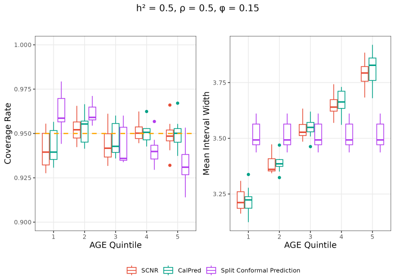
$h^2 = 0.5, \rho = 0.5, \varphi = 0.15$
Coverage Rate
Mean Interval Width
AGE Quintile
SCNR
CalPred
Split Conformal Prediction

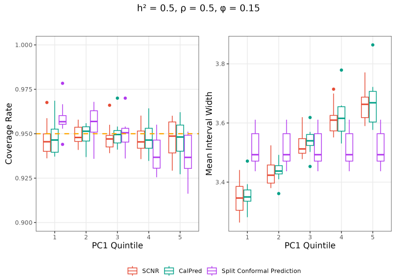
$h^2 = 0.5, \rho = 0.5, \varphi = 0.15$
Coverage Rate
Mean Interval Width
PC1 Quintile
SCNR
CalPred
Split Conformal Prediction

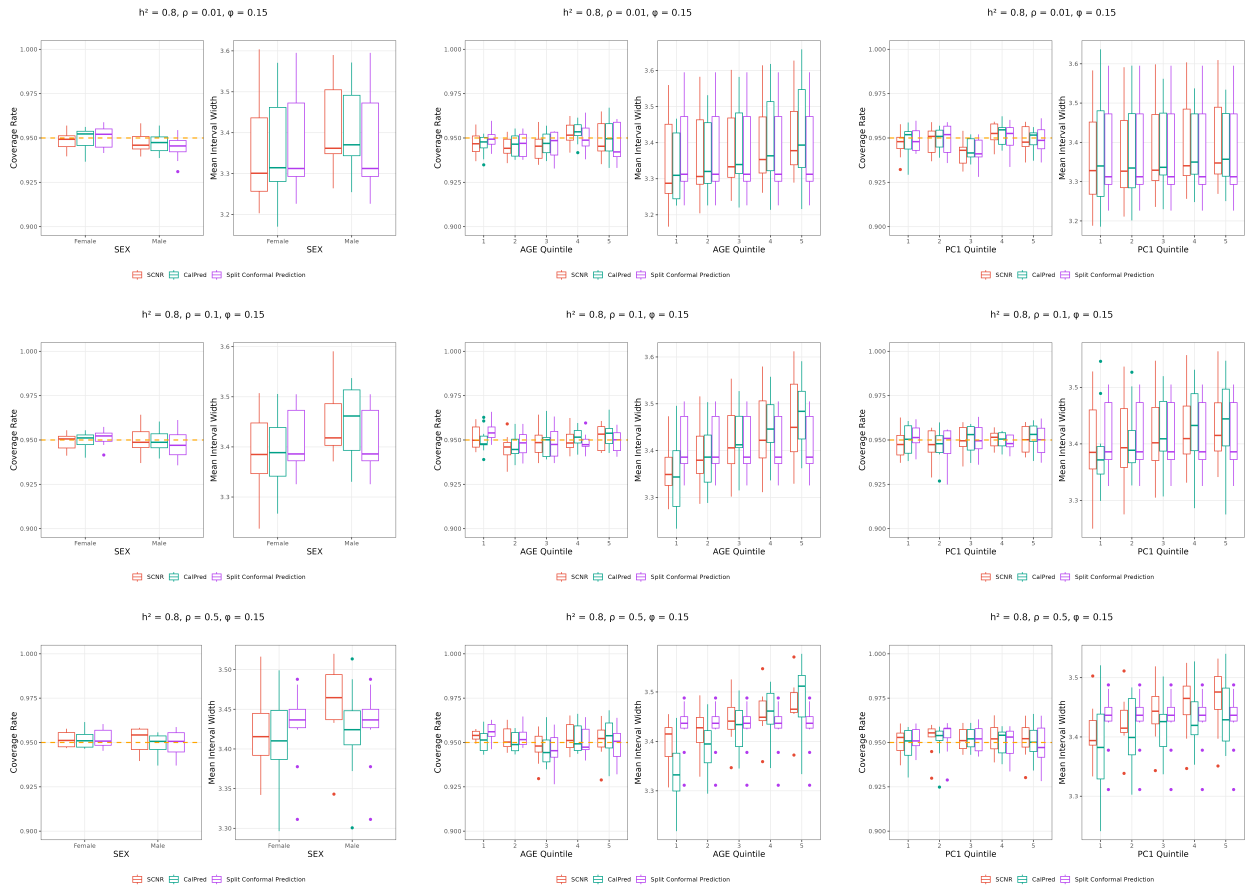


**Supplementary Figure 4. Performance comparison between SCNR, CalPred, and Split Conformal Prediction.** Simulations were performed with a proper training set of $20,000$ individuals and a calibration set of $5,000$ individuals. The prediction intervals for $5,000$ testing individuals were evaluated for each context subgroup (quintiles of age and PC1; female/male for sex). The box plots were constructed based on the 10 simulation replicates for each setting. The dashed orange lines represent the target 0.95 confidence level. The parameters are set to heritability $h^2 = \{0.2, 0.5, 0.8\}$, polygenicity $\rho = \{0.01, 0.1, 0.5\}$, and proportion of variance attributed to covariates $\varphi = 0.15$. Each panel displays the empirical prediction coverage rate (left) and the corresponding mean interval width (right). The panels in each row correspond to one parameter configuration, where **(Left)** subgroups stratified by sex; **(Middle)** subgroups stratified by quintiles of age; **(Right)** subgroups stratified by quintiles of PC1.

### Supplementary Table

**Supplementary Table 1.** Summary of UK Biobank Traits

| **Data Field** | **Trait Name** | $N$ | $N_{\text{Female}}$ | $N_{\text{Male}}$ |
|---|---|---|---|---|
| 21001 | Body Mass Index (BMI) | 313,623 | 168,484 | 145,139 |
| 30780 | Low-Density Lipoprotein (LDL) Cholesterol | 299,388 | 160,740 | 138,648 |
| 30760 | High-Density Lipoprotein (HDL) Cholesterol | 274,597 | 146,293 | 128,304 |
